# Predicting the Effectiveness of Covid-19 Vaccines from SARS-CoV-2 Variants Neutralisation Data

**DOI:** 10.1101/2021.09.06.21263160

**Authors:** Oleg Volkov, Svetlana Borozdenkova, Alexander Gray

## Abstract

Rapid and accurate prediction of Covid-19 vaccine effectiveness is crucial to response against SARS-CoV-2 variants of concern. Despite intensive research, several prediction tasks are not well supported, such as predicting effectiveness of partial vaccination, of vaccine boosters and in vaccinated subpopulations. This paper introduces a novel predictive framework to accommodate such tasks and improve prediction accuracy. It was developed for predicting the symptomatic effectiveness of the BNT162b2 (Comirnaty) and ChAdOx1 nCoV-19 (Vaxzevria) vaccines but could apply to other vaccines and effectiveness types. Direct prediction within the framework uses levels of vaccine-induced neutralising antibodies against SARS-CoV-2 variants to fit efficacy and effectiveness estimates from studies with a given vaccine. Indirect prediction uses a model fitted for one vaccine to predict the effectiveness of another. The directly predicted effectiveness of Comirnaty against the Delta variant was 44.8% (22, 69) after one and 84.6% (64, 97) after two doses, which is close to 45.6% and 85.5%, the average estimates from effectiveness studies with the vaccine. The corresponding direct predictions for Vaxzevria were 41.6% (18, 68) and 63.2% (37, 86); and the indirect predictions, from the model fitted to Comirnaty data, were 45.5% (23, 70) and 61.2% (37, 83). Both sets of predictions are comparable to the average estimates, 42.5% and 66.3%, from effectiveness studies with Vaxzevria. Further results are presented on age subgroups; prediction biases and their mitigation; and implications for vaccination policies.

---

Predictive frameworks based on correlates of immune protection can be a game changer in the fight against the Covid-19 pandemic. They would allow prediction of vaccine efficacy and effectiveness from levels of immune response, such as antibody levels. Unlike with months-long clinical trials and observational studies, the requisite data can be obtained rapidly and efficiently from blood sampling and automated in-vitro studies. Life-critical questions What are the optimal doses and timing of vaccine boosters in a population? How likely is a new variant to evade vaccine immunity? — would be answered more quickly. Despite immense research efforts, the frameworks proposed for Covid-19 vaccines are yet to answer several of such crucial questions.

In developing or evaluating such frameworks, the following aspects could matter:

**Ease of implementation** Additional large human trials are not required. Existing in-vitro technologies are sufficient or easily adapted. Modelling and software tools are simple to use and adapt by practitioners.

**Wide scope** Crucial prediction tasks are covered, including those to support regimen modifications of established vaccines; accelerated development of new vaccines; tailored policies for population subgroups; and risk assessment for emerging variants.

**Validity and accuracy** Predictions within a specific task are relatively unbiased and precise.

Improving upon these aspects could prevent deaths, and there is still a potential for improvement (Tregoning et al., 2021). Given the gravity and complexity of the pandemic, a one-size-fits-all framework may be elusive, and tailored approaches more suitable.

One crucial framework was developed within a purpose-designed trial with a subgroup of vaccinated and placebo participants in the Moderna mRNA-1273 clinical trials (Gilbert et al., 2021). Several immune correlates were established based on symptomatic Covid-19 cases in this subgroup, and rigorous causality analyses were performed. Although large in scope, this study did not cover all practically relevant tasks. As pointed out in Gilbert et al. (2021), it did not consider virus variants, predictions after one dose, or predictions for other SARS-CoV-2 vaccines. While the study used pseudovirus neutralisation assays, evidence has been accumulating that live SARS-CoV-2 neutralisation assays could be more informative (Lu et al., 2021).

A similar framework was developed within ChAdOx1 nCoV-19 (Vaxzevria) vaccine trials (Feng et al., 2021). Several correlates of symptomatic effectiveness of the vaccine were identified, including neutralising titres in live-virus neutralisation studies. Estimates were obtained for immune marker values associated with 80% and other levels of symptomatic efficacy. However, the uncertainty of the estimates was high, and most confidence intervals could not be computed (cf. Table 2 of Feng et al., 2021). Correlates were established for five primary symptoms of Covid-19 but not for other symptoms of the disease. Efficacy prediction at high levels of live neutralisation titres was rather uncertain (cf. Figure 4d of the paper). The study did not investigate protection after a single dose, against variants other than B.1.177 and B.1.1.7, for alternative vaccines and in population subgroups (Feng et al., 2021).

Two groundbreaking frameworks were proposed in Earle et al. (2021) and Khoury et al. (2021), with the latter extended in Cromer et al. (2021). The efficacy in Phase 3 vaccine trials was predicted by using live SARS-CoV-2 neutralising titres from vaccinated volunteers in Phase 1/2 immunogenicity studies. The strengths of the frameworks include combining data across clinical trials and neutralisation studies for different vaccines. To overcome assay disparity between studies, vaccine-induced titres were normalised by mean titres from convalescent plasma, used as a reference in a given study.

This normalisation, however, has trade-offs. For instance, studies without convalescent samples cannot be included. It also adds an extra distribution (of convalescent titres) and hence increases variability. It could introduce biases via various mechanisms. For instance, unvaccinated younger Covid patients could produce lower neutralising titres than older patients would, perhaps due to less severe disease (Li et al., 2021). As the opposite occurs in vaccinated people, this could bias predictions, particularly for age subgroups.

Other potential sources of error must be heeded when applying the frameworks in Earle et al. (2021) and Khoury et al. (2021). For instance, pooling data across neutralisation studies can introduce errors due to different types of neutralisation measured (e.g., *PRNT*_80_ versus *MN IC*_50_; Khoury et al. 2021), and due to other types of variability between laboratories. Differences among vaccines and particularly vaccine platforms (e.g., mRNA versus adenovirus vector) could bias predictions. Error can also be due to incorrect inputs into the modelling. Indeed, convalescent samples used may be unrepresentative of wider convalescent populations. Vaccinated individuals in Phase 1/2 immunogenicity studies may not generalise to wider populations in Phase 3 trials and beyond. These issues are exacerbated by typically small sizes of early studies, such as 15 vaccinated and 3 convalescent subjects in the Moderna study (Jackson et al., 2020) assumed in Khoury et al. (2021). Unless such errors are addressed, a model could fit current data well but produce biased predictions.

Although the frameworks in Earle et al. (2021) and Khoury et al. (2021) accurately predicted vaccine efficacy, they could still be augmented for some applications. Both frameworks focused on full vaccination (e.g., with two doses of mRNA vaccines), whereas prediction after one dose could be also important. Their predictions may become increasingly uncertain for emerging SARS-CoV-2 variants, since the assumed Phase 1/2 neutralisation studies were done with the virus Wild Type, whereas Phase 3 efficacy data were collected already with its successors. For example, the predominant genotype during the Phase 3 Pfizer-BioNTech BNT162b2 (Comirnaty) vaccine trials was D614G (Korber et al., 2020), which could markedly decrease neutralisation compared with the Wild Type (e.g., by 2.3 folds according to Wall et al. 2021b). Delta and successor variants would be further steps away from the Wild Type. Another potential task is prediction for age subgroups, which was not addressed by the frameworks.

In this paper we present a novel prediction framework for SARS-CoV-2 vaccines, which was built upon Earle et al. (2021) and particularly Khoury et al. (2021). As it utilised neutralisation data from a single large-scale study (described in Wall et al., 2021b,a), normalisation by convalescent titres was not required, thus eliminating its associated error, along with error due to inter-laboratory variation.

Live-virus neutralising titres were established as correlates of protection in the trial reported in Feng et al. (2021) and via modelling in Khoury et al. (2021) and Earle et al. (2021). Similarly to the data assumed in Khoury et al. (2021) and Earle et al. (2021), virus neutralisation and vaccine effectiveness data assumed in this paper were obtained in different groups of subjects. Generalisability of neutralisation study populations to wider populations is therefore a key concern. As steps towards addressing it, we investigated potential impacts of population differences numerically and suggested mitigation strategies.

The proposed framework could enhance prediction accuracy and complement the existing frameworks in applications. Its focus is on vaccine effectiveness after full and partial vaccination, in age subgroups, and against SARS-CoV-2 variants. It could also help predict efficacy for clinical trials with an updated vaccine. Although the framework was developed for the Comirnaty and Vaxzevria vaccines, it could apply to other vaccines with sufficient neutralisation and effectiveness, or efficacy, data. The required modelling and software are simple to apply and potentially adapt.

The framework has limitations and makes several trade-offs. For instance, like Earle et al. (2021) and Khoury et al. (2021) it did not investigate causality and could only provide indirect evidence for correlates of protection. Its implementation does not yet utilise data across neutralisation studies. Some tasks — such as predictions for new vaccines under development — could be addressed by extending the framework with elements of other frameworks.

We next present results from direct modelling for Comirnaty, which is based model fitting to data on this vaccine. This is followed by results from similar direct modelling for Vaxzevria. We also present results from indirect modelling: predictions of Vaxzevria effectiveness made from a model fitted to Comirnaty data. The appendix presents detailed methodology; an investigation of Age and other covariates for neutralising titres; and an assessment of biases due to population differences along with mitigation strategies. The assumed neutralisation and effectiveness studies with the vaccines are summarised.

## 1 Results

### 1.1 Modelling for the Comirnaty vaccine

To model symptomatic effectiveness we used its estimates from the Pfizer-BioNTech Phase 3 trial (FDA, 2020; Polack et al., 2020) and from seven observational studies with the Comirnaty vaccine post-authorisation. We included studies that either reported symptomatic effectiveness against specific variants, or in which the single dominant variant (accounting for 80% or more cases) could be established from study dates and locations. These studies were conducted in Canada (Nasreen et al., 2021), France (Charmet et al., 2021), Scotland (Sheikh et al., 2021) and two in England (Bernal et al., 2021; Pouwels et al., 2021) and Israel (Haas et al., 2021; Mor et al., 2021) each. (See the Methods Supplement for more details.)

The correlates of immune protection were based on data obtained at the Crick Institute within the ongoing Legacy Study (Wall et al., 2021b). The study ran one cohort with *N* = 149 healthcare workers in the UK who had had a single dose of the Comirnaty vaccine, and another cohort, with *N* = 159, who had had both doses. Plasma collected from the participants was tested in automated live-virus neutralisation assays against the D614G, B.1.1.7 (Alpha), B.1.351 (Beta) and B.1.167.2 (Delta) variants. The features of the resulting neutralisation titres are summarised in the Supplement.

Each distribution of neutralisation titres per dose and variant was paired with the effectiveness observations for the dose and variant. These data were fitted using the population protection function (2), considered in the Supplement, which is common in correlates of protection research (Khoury et al., 2021; Dunning, 2006; Nauta et al., 2009).

Unlike Khoury et al. (2021), who assumed a Normal distribution for vaccine efficacy, we assumed a beta distribution for vaccine effectiveness. The latter distribution is more suitable for outcomes on a bounded interval — such as between 0% and 100% in our case — than a normal distribution with an unbounded domain. Given our assumption, we fit the population model using nonlinear beta regression. To the best of our knowledge beta regression modelling has not been applied in vaccine research. (Modelling and methodology details are in the Supplement.)

The model fit is plotted in Figure 1 against the geometric mean titre (GMT) per variant sample. (As our model function inputs empirical distributions, this figure is a simplified 2D representation of a multi-dimensional problem. Caution is therefore required in interpretation. More details and an approximation to the model are considered in Appendix B.)

**Figure 1:**
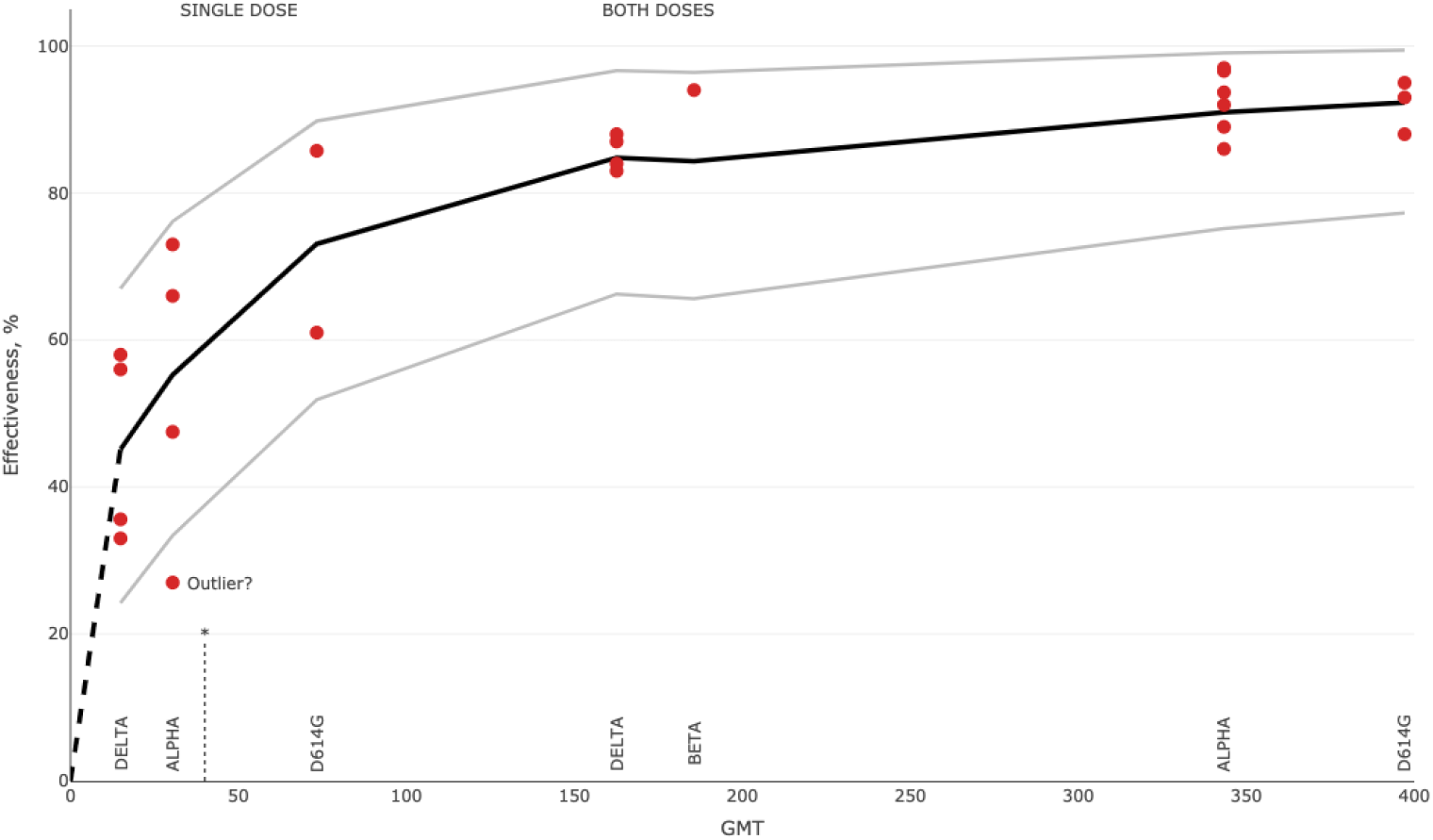
Model fitting observed symptomatic effectiveness of Comirnaty. Each group of observations corresponds to the variant indicated and is plotted against the geometric mean titre from the corresponding Crick sample. The 95% prediction bands are shown. The dotted line indicates the lower quantification threshold at *GMT* = 40. The dashed line extends the model fit beyond the data range towards zero.

Figure 1 suggests a substantial nonlinear association between neutralising titres from the Crick study and the observations from the effectiveness studies. The association is well represented by the assumed model. This is despite the studies being performed in different countries, at different times post-vaccination and in diverse populations.

The model also characterised the prediction uncertainty well, with 23 out of 24 observations lying within 95% prediction interval. The exception is the single-dose symptomatic effectiveness, in %, of 27(13, 39) against the Alpha variant from the Scotland study in Sheikh et al. (2021). This is a potential outlier, as it is smaller than every symptomatic effectiveness estimate against the Delta variant in the studies considered here. It is also smaller than the effectiveness against infection caused by either the Alpha 38(29, 45) or Delta 30(17, 41) variant in Sheikh et al. (2021). That our model identified this outlier gives an additional reassurance about its quality.

Variability in the effectiveness observations is greater after a single dose and peaks in the region of 50% effectiveness, a pattern echoed by the prediction bands in Figure 1. This is unsurprising, since the uncertainty of a beta random variable is maximal in the middle of its domain. Hence there is additional empirical evidence for assuming a beta distribution.

### 1.2 Predictions for the Comirnaty vaccine

#### Effectiveness in age subgroups

Appendix C explores dependence of neutralising titres on Age and other covariates, and considers potential confounding. The results in the appendix imply that the larger age groups considered in this section — below 35 and 50 and over — were well balanced in the Crick study with respect to potential confounders. (The participants’ ages were roughly between 20 and 70.) This holds for both cohorts, although the single-dose case requires caution due to many titres under the detection threshold; see Figure 12. The other age groups in this section below 30 and 60 and over — were small and relatively unbalanced with respect to the confounders. However, as discussed in Appendix C, confounding is relatively small and could be inconsequential even for these age groups. (Also refer to Tables 1 and 2.)

**Table 1:** Linear regression of log10 titres against Delta in the both-doses cohort

| Coefficient | Estimate | SE | P-value |
| --- | --- | --- | --- |
| Crude regression |  |  |  |
| Intercept | 2.5608 | 0.1133 | 0.0000 |
| Age, years | -0.0081 | 0.0025 | 0.0017 |
| Multivariate regression |  |  |  |
| Intercept | 2.4275 | 0.1252 | 0.0000 |
| Age, years | -0.0089 | 0.0024 | 0.0002 |
| Dosing Interval > 40 days | 0.2169 | 0.0561 | 0.0002 |
| Prior Covid | 0.1857 | 0.0576 | 0.0015 |
| Days Since Jab 2 | -0.0007 | 0.0024 | 0.7628 |
| Sex (Male) | 0.0102 | 0.0555 | 0.8551 |

**Table 2:** Linear regression of log10 titres against Delta in the single-dose cohort

| Coefficient | Estimate | SE | P-value |
| --- | --- | --- | --- |
| Crude regression |  |  |  |
| Intercept | 1.4805 | 0.1758 | 0.0000 |
| Age, years | -0.0072 | 0.0040 | 0.0709 |
| Multivariate regression |  |  |  |
| Intercept | 1.4640 | 0.2090 | 0.0000 |
| Age, years | -0.0080 | 0.0040 | 0.0485 |
| Prior Covid | 0.2825 | 0.1083 | 0.0099 |
| Sex (Male) | 0.1158 | 0.1085 | 0.2873 |
| Days Since Jab 1 | -0.0017 | 0.0045 | 0.7077 |

The differences in neutralising titres between participants under 35 with those 50 and over are illustrated in Figure 13. In almost all cases, the younger group had 1.6-2.3 times higher GMT than the older group. The model fitted using all-age neutralisation data was applied to predict effectiveness against the Delta variant in selected age groups; see Figure 2.

**Figure 2:**
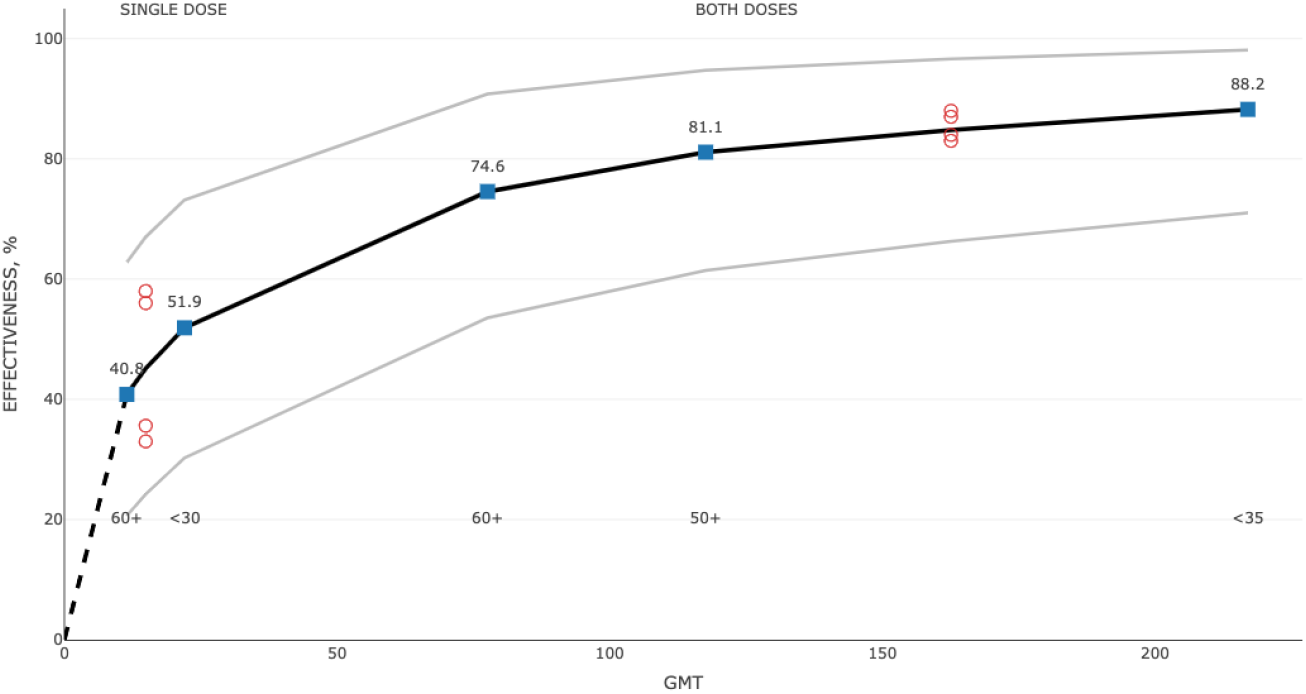
Predicted effectiveness of Comirnaty against the Delta variant for the indicated age groups. The model fit, the same as in Figure 1, for all ages was used for this prediction. The effectiveness observations were against Delta in all-age groups.

#### Effectiveness against variants

We first applied our framework to predict effectiveness against the Delta variant. The model was fitted using neutralisation and effectiveness data on the assumed variants except Delta. Next, the neutralisation titres against Delta were input into the fitted model to predict effectiveness against this variant. These predictions were compared with estimates of symptomatic effectiveness against the variant in Nasreen et al. (2021), Sheikh et al. (2021), Bernal et al. (2021) and Pouwels et al. (2021), detailed in Appendix E. The model fit and observations are plotted in Figure 3.

**Figure 3:**
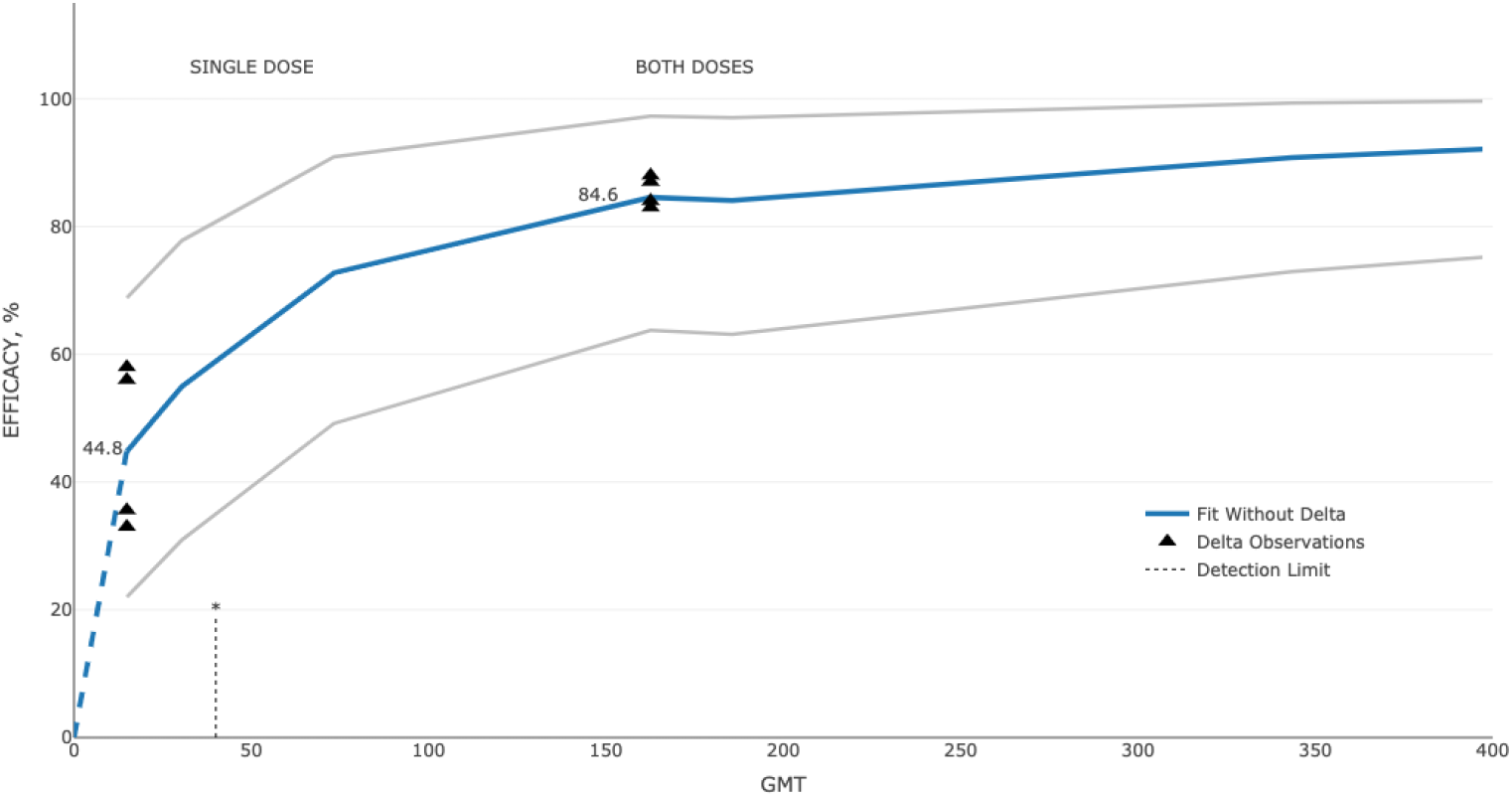
Predicted effectiveness of Comirnaty against the Delta variant. The model was fitted using the effectiveness and neutralisation data on all the variants except the indicated variant. Prediction was evaluated at the neutralisation titres of the variant.

Similar predictions were done for D614G and Alpha (but not for Beta, as only one effectiveness estimate was available.) The results are illustrated in Figure 4. The predictions were remarkably close to the corresponding mean effectiveness values from effectiveness studies. For instance, for the Delta variant there was an underprediction of only 0.8% after one dose, and of only 0.9% after two doses. The largest error — an overprediction of 3% — was for the Alpha variant after one dose.

**Figure 4:**
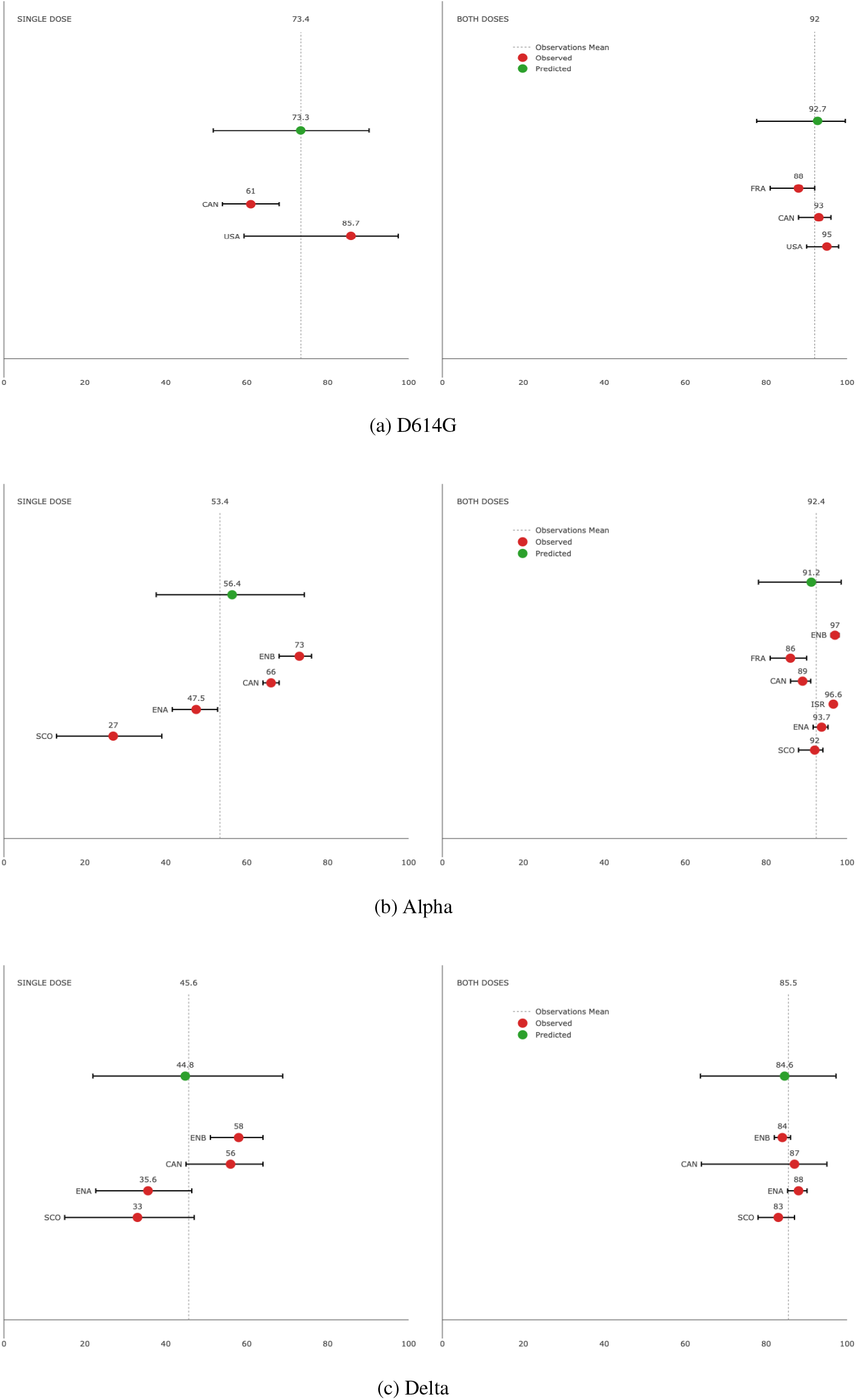
Predicted effectiveness of Comirnaty against a variant. Estimates from effectiveness studies are shown versus predictions from a model fitted without the variant indicated. 95% prediction bands and means of observations are shown. Study countries are indicated, with ENA and ENB standing for England study A and B, respectively.

The prediction uncertainty appears realistic; see Figure 4. In all but one cases the 95% prediction interval included the corresponding estimates from the effectiveness studies. The exception was the effectiveness estimate after one dose from the Scotland study (Sheikh et al., 2021). (This estimate is a suspected outlier; see Section 1.1.) The prediction bands largely included the confidence intervals of effectiveness estimates.

Notably, prediction is more difficult after a single dose than after both doses:

- There is larger uncertainty in effectiveness values around 50%, as evidenced by a greater spread of effectiveness observations.
- The neutralisation titres after one dose could lie outside the neutralisation data range used in the fitting of the other variants. This was the case for the Delta variant, whose GMT was 13.3.
- As was also the case for Delta, the titres could lie well below the lower detection threshold of 40; see Figure 3. Left censoring could be particularly impactful, and predictions sensitive to the assumed nominal values, 10 and 5, representing weak and absent neutralisation signals, respectively (see the Supplement for details).
- As the model function is relatively steep at small titres, the impact of differences between true and assumed titres is magnified disproportionately.

As more individuals get fully vaccinated, prediction uncertainty after one dose becomes less important in practice.

### 1.3 Modelling and prediction for the Vaxzevria vaccine

The required neutralisation titres were obtained from the Vaxzevria stream of the Crick Legacy study (Wall et al., 2021a). This stream included a single-dose cohort with 50 and a two-dose cohort with 63 healthcare workers vaccinated with Vaxzevria. Live virus neutralisation was investigated against the same variants as in the Comirnaty stream.

We included estimates of symptomatic effectiveness of Vaxzevria from Emary et al. (2021), based on the vaccine clinical trial (Ramasamy et al., 2020), and from four observational studies with the vaccine post-authorisation: one in Canada (Nasreen et al., 2021); two in England (Bernal et al., 2021; Pouwels et al., 2021); and one in Scotland (Sheikh et al., 2021). Altogether there were 17 observations for D614G, Alpha and Delta variants in all-age populations after one or two doses.

We first modelled effectiveness of Vaxzevria directly, by fitting these observations to neutralisation data for this vaccine. Note the higher underlying uncertainty for Vaxzevria than for Comirnaty due to the cohorts being smaller and more skewed towards younger ages, and due to fewer effectiveness observations (17 v 24). We also applied indirect modelling by using the model fitted to Comirnaty data to predict the effectiveness of Vaxzevria.

The two model fits are compared in Figure 5, where they are evaluated at Vaxzevria neutralisation titres. Panel A of the figure suggests that the fits are similar in shape and relatively close: the difference between the Vaxzevria and Comirnaty fits is between -3.3% and 6.1%. Panel A indicates that the Vaxzevria effectiveness estimate of 97% against Alpha after two doses is a potential outlier. This estimate, given in Pouwels et al. (2021), was based on self-reported Covid-19 symptoms and involved relatively few (56) vaccinated people; see Figure S5 of that paper. To the best of our knowledge, this estimate exceeds every other published estimate of symptomatic effectiveness for Vaxzevria. (Refer, for example, to Figure 3 of Imai et al. (2021).) Panel B of Figure 5 shows a model fit for Vaxzevria with this estimate removed. This fit is noticeably closer to the Comirnaty fit, with the difference between the fits being between -1.8% and 1.3%.

**Figure 5:**
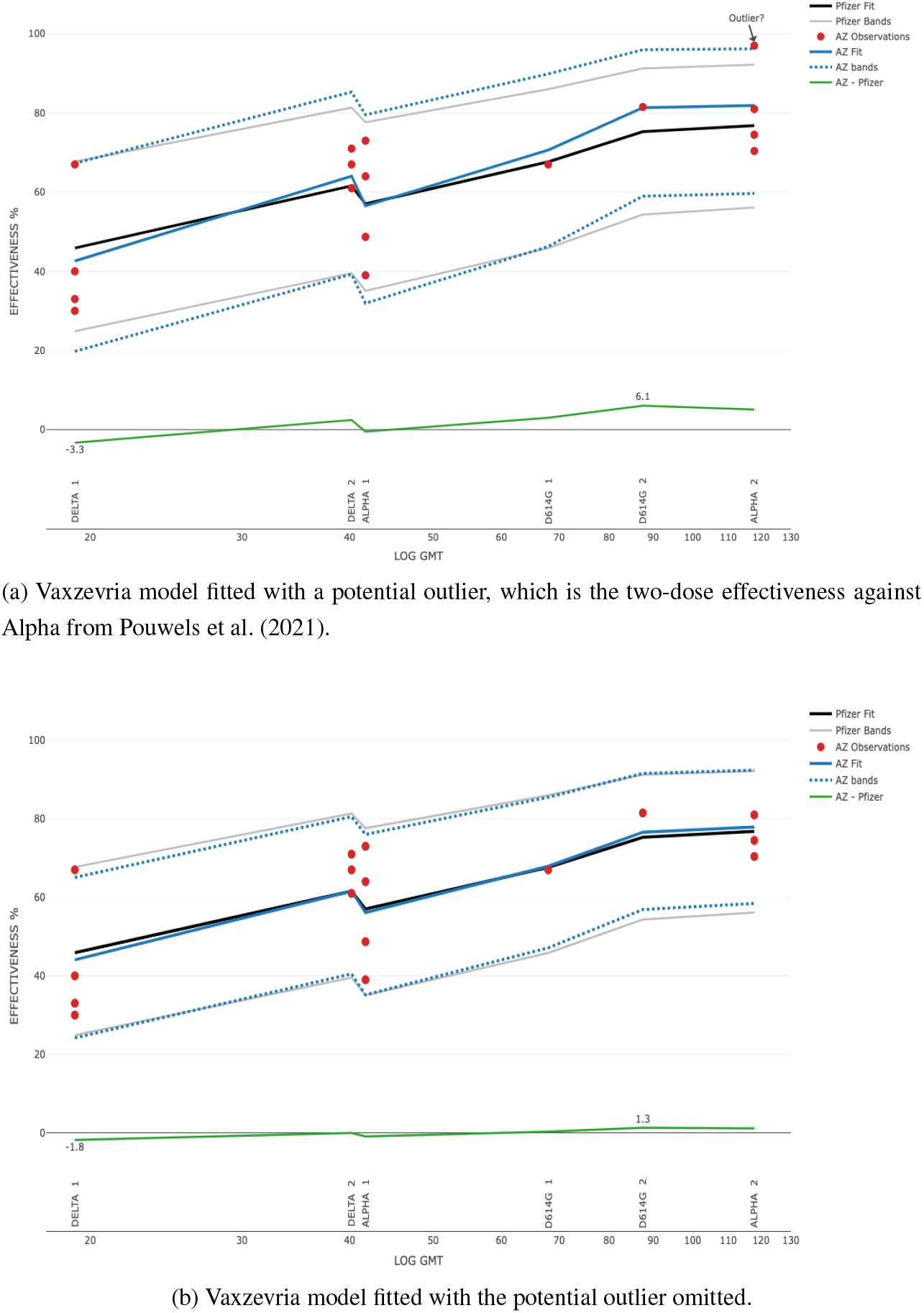
Comirnaty model fit is plotted against log titres, observations and model fit for Vaxzevria. The largest differences between the models are indicated.

#### Prediction for specific variants

We applied the two modelling approaches — direct fitting to Vaxzevria data and the use of Comirnaty fit — to predict symptomatic effectiveness of Vaxzevria against the Alpha and Delta variants. (D614G was not assessed as there were only two effectiveness observations for it.) The fits were obtained without the data on the variant under consideration. Each fitted model was evaluated at the Vaxzevria neutralising titres with the variant; see Figure 6. Direct fitting to Vaxzevria data including the potential outlier produced closer predictions than the fitting to Comirnaty data. However, the differences between the predictions were relatively small: about 4% at most. When the outlier was omitted, predictions from the Vaxzevria fit were very similar to those from the Comirnaty fit; see Panel B of Figure 6.

**Figure 6:**
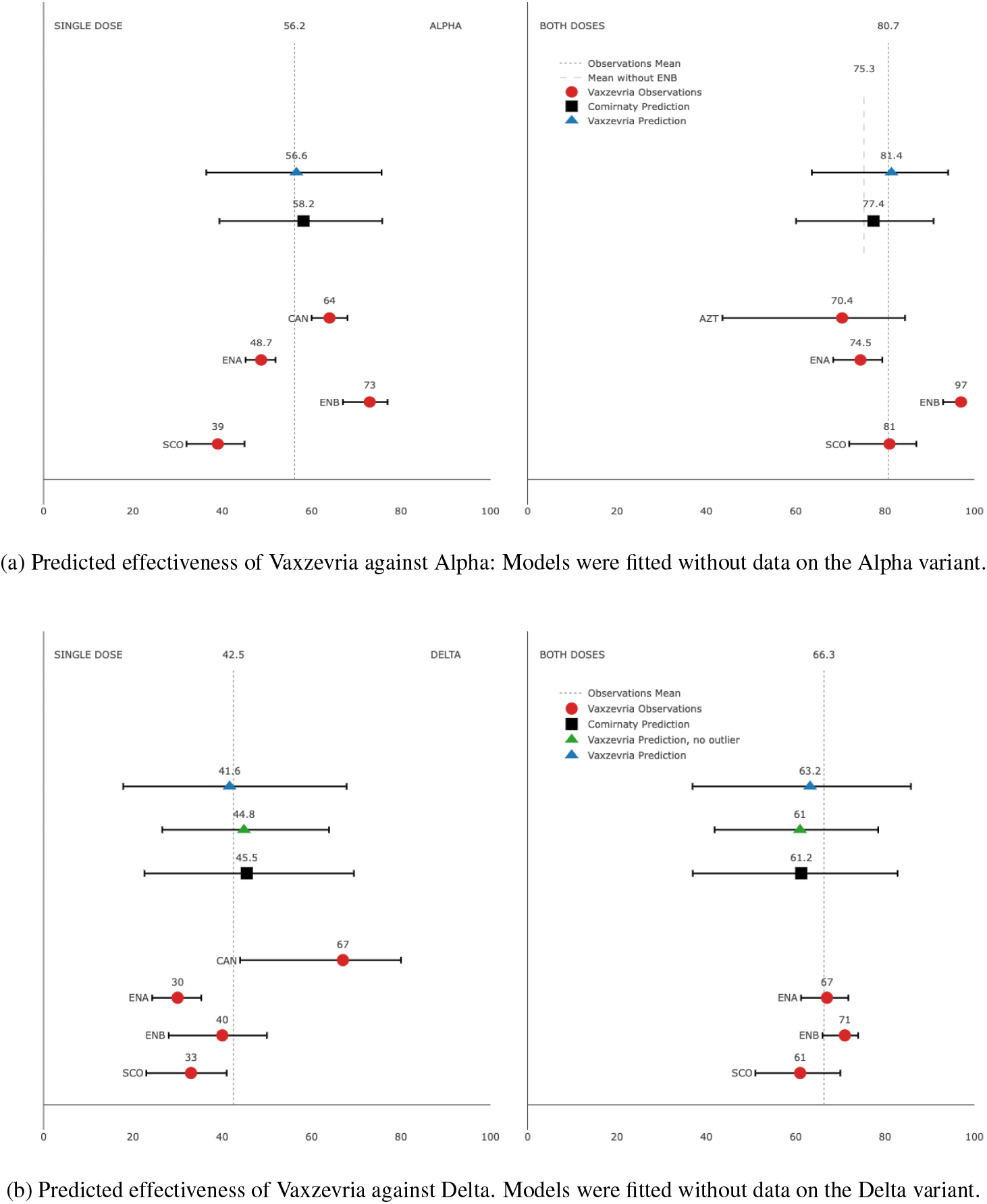
Predicted symptomatic effectiveness of Vaxzevria against SARS-CoV-2 variants. Comirnaty prediction was based on Comirnaty data; Vaxzevria prediction on Vaxzevria data. Effectiveness observations and 95% confidence intervals are shown for the Vaxzevria. Study countries are indicated, with ENA and ENB standing for England study A and B, respectively. AZT stands for the original AZ/Oxford Phase 3 vaccine trial.

## 2 Discussion

The proposed framework produced accurate predictions of vaccine effectiveness against SARS-CoV-2 variants after one and two vaccine doses. This is despite potentially large differences between the Crick study population and the populations in the effectiveness studies, as well as the likely disparities between effectiveness definitions in different studies, and a generally high uncertainty after one dose. The predictions of Vaxzevria effectiveness from a Comirnaty model fit were close to predictions from a directly fitted Vaxzevria model. This is despite the use of data from two different vaccines (and vaccine platforms) and four different dose cohorts in neutralisation studies.

Our modelling established a strong nonlinear association between the neutralisation titre distributions and the effectiveness observations assumed. Although we did not examine causality, the high accuracy of predictions gave an indirect support for the use of neutralising titres as correlates of protection.

We pinpointed potential biases due to population differences between neutralisation and effectiveness studies. Effectiveness predictions could benefit from population adjustments, such as those considered in Section D.2, as well as from cautious prediction, considered in Section D.5.

### Future work

It can be beneficial to incorporate further adjustments for populations into our framework. Our modelling could be additionally validated with detailed data from past and future effectiveness studies. Modelling refinements could include incorporation of titre measurement errors; additional covariates; between-study variability; imputation and other methods concerned with censoring.

A valuable extension of our framework would be to combine data across neutralisation studies, perhaps by adapting the methods in Khoury et al. (2021) and Earle et al. (2021) and using data conversion to WHO-proposed standards for SARS-CoV-2 neutralisation data. A relatively straightforward extension is to accommodate various types of vaccine effectiveness and efficacy; and also effectiveness against a mix of SARS-CoV-2 variants. Furthermore, our framework could be validated by comparing its predictions with those from the frameworks in Earle et al. (2021), Khoury et al. (2021), Gilbert et al. (2021) and Feng et al. (2021). It could also be valuable to incorporate elements from these frameworks, for example, the correlates of protection established in Feng et al. (2021) for the Alpha variant.

### 2.1 Policy implications

#### Monitoring neutralisation titres

Our modelling supports conclusions in Wall et al. (2021a) about the benefits of monitoring neutralisation titres in populations. Indeed, the Crick study already enabled accurate predictions of vaccine effectiveness for the three SARS-CoV-2 variants. It could be representative of larger populations of healthcare workers and of general working-age populations. Any follow-up data from the study, for instance, on waning immunity or on response to a third booster could be highly informative and valuable for policy decisions.

Expanding the Crick study or running additional cohorts and studies of SARS-CoV-2 variants neutralisation could bring immense further benefits. These could include early warnings of waning immunity in subpopulations, optimisation of booster schedules and rapid identification of immunity-evading variants. Likewise, studies could predict vaccine effectiveness against successors of existing variants of concern, such as Delta Plus.

#### Human challenge studies

It could be possible to validate correlates of protection models in human challenge studies with a vaccine. To emulate lower antibody levels in older populations, and to provide information on dose reduction potential, a lower vaccine dose could be administered to the study participants (who are usually aged between 18 and 30). As effectiveness could be modelled across variants, safer challenges with more benign variants, such as D614G, may be sufficient.

#### Effectiveness regions

Our modelling suggests three regions for vaccine effectiveness, which could be relevant to decision-making. These regions are summarised for the Comirnaty vaccine.

**Flat region** includes effectiveness against every variant after two doses in the all-age population (e.g., Figure 1). This region allows maximum flexibility for policy-making, since moderate changes in neutralising titres will have only a small impact on effectiveness. For example, Payne et al. (2021) reported greater neutralisation for the Comirnaty vaccine with an eight-week dosing interval than with three-to-five week intervals used in the clinical trials. (This also appears to be the case for the Crick study; see Figure 10.) However, a longer interval could expose partially vaccinated persons to higher infection risk, since single-dose protection is lower. Within a flat region, either longer or shorter intervals could be chosen on epidemiological grounds, without risking vaccine effectiveness.

There is also little effectiveness cost from reduced vaccine doses. Policy-makers could confidently decide on dose reduction so as to address supply shortages, high costs and dose-dependent toxicities. This could also free up doses for vulnerable populations worldwide.

Over time, waning vaccine protection could drive neutralisation titres– particularly in the elderly and vulnerable — outside the flat region, especially for the Delta variant. Still, a flat region could be reached again after another booster (i.e., a third dose). Boosting with reduced doses would have little impact on effectiveness, compared to full-dose boosters. Furthermore, a Comirnaty vaccine update for the Delta variant or a new universal vaccine could probably be administered in smaller doses than the current doses.

**Steep region** corresponds, for example, to effectiveness against Alpha or Delta after a single dose. This region gives less leeway for vaccination policies: For instance, reducing the dose may cause a relatively steep drop in effectiveness. First-dose reduction could still be beneficial, if implemented with due caution. For instance, if the pandemic is under control by non-pharmaceutical measures, reduced first doses could accelerate reaching full vaccination and protection of a country’s population.

**Intermediate region** is located between the other two regions. It is exemplified by the single-dose effectiveness against D614G for all ages (Figure 1) and possibly by the two-dose effectiveness against Delta in the elderly (Figure 2). Its distinguishing feature is an increased uncertainty about effectiveness, as reflected by a beta distribution. Policies for an intermediate region could combine those for the other two, while accounting for the extra uncertainty.

These regions and their interpretation would be different for vaccine effectiveness against hospitalisation and death. We note that predictions within a flat region are less sensitive to modelling assumptions and population differences. Conversely, predictions within a steep region could be more sensitive due to the steepness of the model function and proximity to the lower detection limit.

##### Age differences

We found considerable differences in virus neutralisation and vaccine effectiveness between age groups, which could warrant age-tailored vaccination policies. For example, the group of twice-vaccinated participants under 35 had a 1.9-fold (1.4, 2.6) GMT against the Delta variant versus the GMT in the 50 and older group, and were inside the flat region; see Figure 2. With the Delta variant, the twice-vaccinated 60+ age group appears to be within a region of reduced effectiveness and increased uncertainty. The situation could be worse for wider elderly populations, since the Crick study participants were under 70 and healthy enough to be in employment.

##### Dosing strategies

Our modelling suggests a compromise dose-sparing strategy to provide a full first dose but reduce the initial booster (second dose) in younger age groups, and also the second booster (third dose) in all age groups. Reduced doses in younger groups may be justified — even despite Delta — not only to expand vaccine coverage but also to reduce toxicity and vaccine hesitancy, which are prevalent in these groups. This could particularly make sense for those under 18.

##### Estimates from effectiveness studies

Our framework could give insights into various aspects of pandemic policy. One example is a surprisingly low estimate, of 64%, for symptomatic effectiveness of the Comirnaty vaccine against Delta (Israel Ministry of Health, 2021; Zimmer, 2021). Our modelling implies that this estimate might not be completely surprising for older populations (see Figure 2), given that vaccine effectiveness wanes over time. Another point is that as an intermediate effectiveness region is approached, upcoming effectiveness estimates can become increasingly uncertain. Hence, vaccine boosting decisions may be suboptimal unless based on several effectiveness studies.

##### Addressing current and future variants

The Delta variant already pushes vaccinated populations towards diminished effectiveness, particularly the elderly and vulnerable. (This is also evident from Khoury et al. (2021) and Wall et al. (2021b).) A new variant reducing neutralising titres even further could push the populations even more into the danger zone. Measures such as expanding worldwide vaccine coverage to reduce mutations and variants emergence, as well as possible updating of existing vaccines are needed.

## Data Availability

All data and R code used for this paper will be freely available in October 2021.

https://github.com/spockoyno/pfizer_biontech_vaccine_paper

## Data Availability

All data and R code used for this paper will be freely available in October 2021.

https://github.com/spockoyno/pfizer_biontech_vaccine_paper

## Data Availability

All data and R code used for this paper will be freely available in October 2021.

https://github.com/spockoyno/pfizer_biontech_vaccine_paper

## Data availability

All data and R code used for this paper will be freely available upon publication from the paper’s GitHub repository

## Acknowledgements

O.V. is grateful to Rosemary Bailey, Ian Bartlett and Andrew Langley for their support and encouragement during this research. The authors thank Rebecca Lodwick, Barbara Bogacka and Sheila Bird for their suggestions which substantially improved the paper. We are grateful to the authors of Gilbert et al. (2021), Khoury et al. (2021) and Wall et al. (2021b) for making their data and programming code publicly available, which greatly facilitated our research. We also thank David L. Bauer for providing additional data from the Crick Legacy Study.

## A Methods

### A.1 Neutralisation Titres

The explanatory variable in our modelling was the antibody titres attaining 50% neutralisation of SARS-CoV-2 variants after one or two vaccine doses. The data were obtained at the Crick Institute within the ongoing Legacy study, which run a stream with Comirnaty-vaccinated (Wall et al., 2021b) and a stream with Vaxzevria-vaccinated (Wall et al., 2021a) healthcare workers.

The Comirnaty stream included a one-dose cohort with 149 participants; a two-dose cohort with 159; and 250 unique participants in total (there was some overlap between the cohorts). The Vaxzevria stream included a one-dose cohort with 50 participants; a two-dose cohort with 63; and 106 unique participants in total.

Samples of participants’ plasma were tested in high-throughput automated assays of live-virus neutralisation against the Wild Type, D614G, Alpha, Beta and Delta variants. Anonymised data for both vaccines are available at the GitHub repository for Wall et al. 2021a.

The distributions of log10-transformed titres for Comirnaty are shown in Figure 7. These distributions after the first dose noticeably violate the Normality assumption. The distributions after both doses appear more Normal.

**Figure 7:**
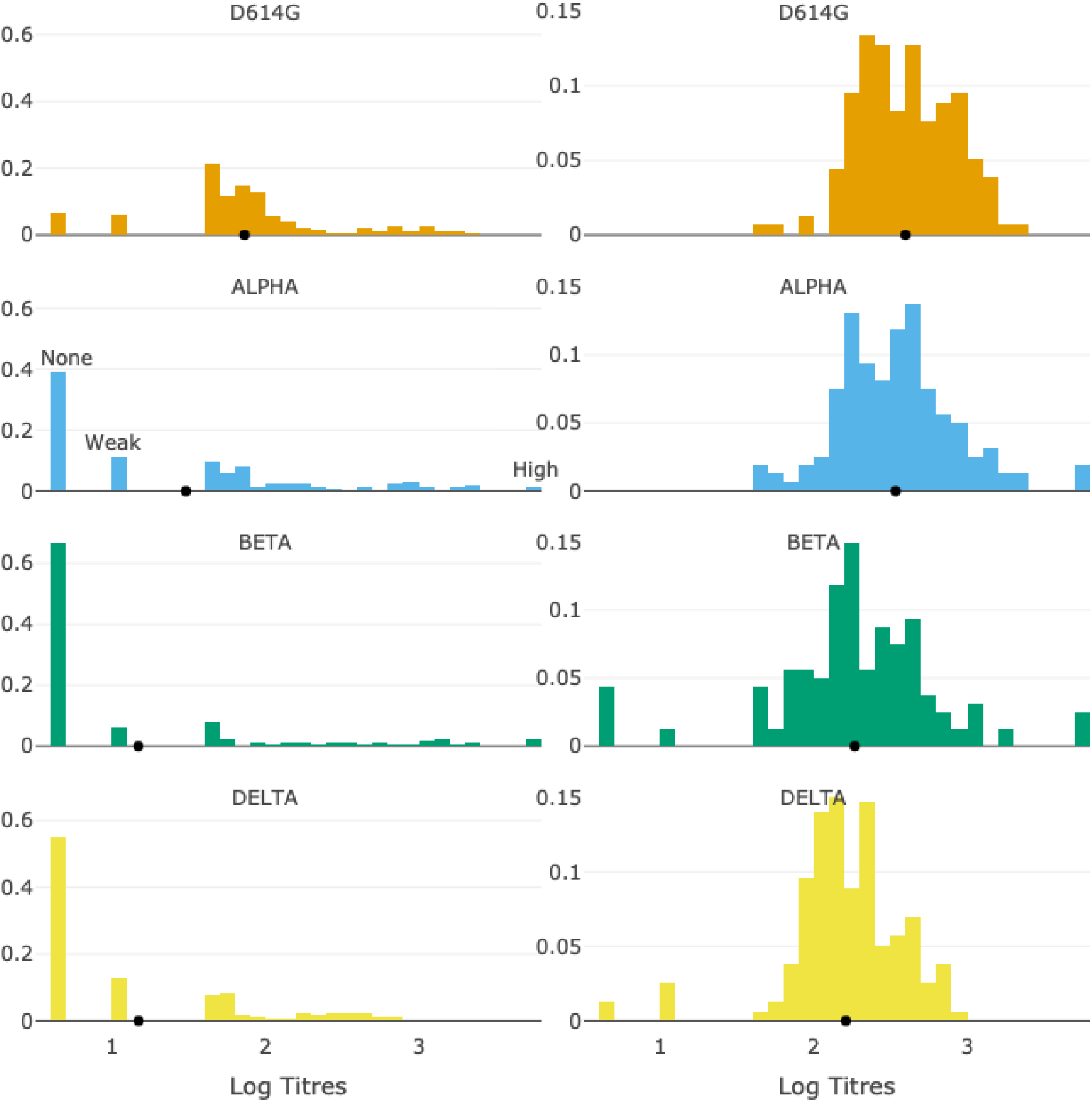
Distributions of log10 titres after one (Left) and two (Right) Comirnaty doses. Detection thresholds are illustrated for Alpha after one dose: “None”, recorded as 5, means no signal; “Weak”, recorded as 10, means a signal below 40; “High”, at 5120, means a signal above 2560. Sources: Wall et al. (2021b).

Any neutralisation signal above the assay quantitative detection limit of 2560 was recorded as a titre of 5120 (Wall et al., 2021b). Any signal below the quantitative detection limit of 40 was recorded as a titre of 10. If no neutralisation signal was observed, a titre of 5 was recorded. These thresholds are illustrated in Figure 7.

### A.2 Protection models

This section draws on Dunning (2006); see also Qin et al. (2007) for a related approach.

#### Individual protection function

The probability of a person with a particular titre being protected against symptomatic Covid-19 was assumed as a scaled logistic function

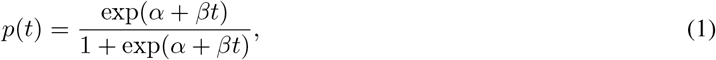

where the neutralising log-titre *t* is the covariate and the constants, *α* and *β*, are the model parameters to be estimated. As is common in vaccine research, we used log_10_-transformed titres. The same function was used in Dunning (2006) and Nauta et al. (2009), and, under a different parametrisation, in Khoury et al. (2021). We assume that this function would have the same parameter values for every individual and across SARS-CoV-2 variants.

#### Population protection function

The proportion protected in a population with *N* people is assumed as

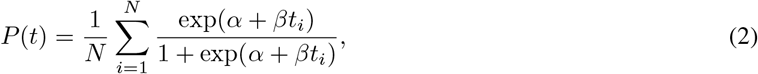

where *t*_*i*_ is the log-titre obtained in person *i* against a particular variant. This function is an expectation over the individual protection functions with the same parameters *α* and *β*. Unlike Khoury et al. (2021), who assumed normal distributions of log-titres, we evaluated this expectation using empirical distributions of log-titres from the Crick study, due to violations of normality and non-standard censoring.

#### Relation to vaccine effectiveness

Following Dunning (2006), we write vaccine efficacy as

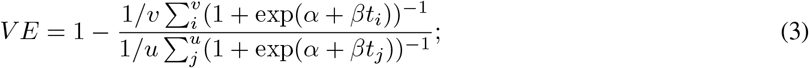

for *v* vaccinated and *u* unvaccinated subjects in a vaccine effectiveness study.

We assume that unvaccinated subjects have negligibly small levels of neutralising antibodies and/or the number of seropositive unvaccinated subjects is negligibly small. Then, unvaccinated subjects’ neutralisation titres would approach zero, and so each log-titre *t*_*j*_ would approach −∞. Hence, each term exp(*α* + *βt*_*j*_) in the denominator of (3) would tend to zero, so that the denominator would approach unity. Therefore, we can write

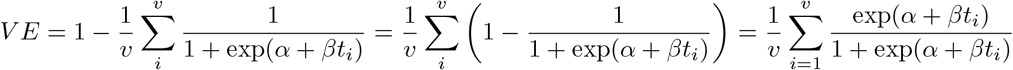

That is, assuming infection-naive unvaccinated subjects, vaccine effectiveness equals the population protection function in (2).

The assumption is unlikely to hold for every vaccine study. However, since prior Covid-19 cases can bias vaccine effectiveness downward (World Health Organisation, 2021, p. 27), study investigators seek to eliminate this bias by excluding such cases from effectiveness analyses. Accordingly, we assume (2) as the model function for (symptomatic) effectiveness of a vaccine.

### A.3 Statistical modelling

#### Modelling the outcome

Observational studies typically estimate vaccine effectiveness by using multi-covariate logistic regression. The resulting estimates can be far from crude estimates based on vaccinated and unvaccinated case ratios in these studies. Therefore, rather than use symptomatic/asymptomatic case counts within a binomial framework, as in Khoury et al. (2021), we fitted the effectiveness estimates directly.

Since these estimates are percentages, they can be treated similarly to probabilities. We therefore used a beta distribution, which is standard for modelling such outcomes. For simplicity, we did not include the endpoints 0% and 100%, which can be accommodated by an extended beta distribution.

#### Beta regression

The model function (2) was fit using nonlinear beta regression (Ferrari and Cribari-Neto, 2004). The corresponding log-likelihood function was maximised numerically with the *optim* procedure in R. To obtain a starting guess for maximisation we fitted the individual protection function (1) to the geometric mean titres (GMTs) per variant/dose combination. Since a logit transformation of (1) is *α* + *βt*, this was done within a generalised linear model (GLM) framework of beta regression. Specifically, the *betareg* package (Cribari-Neto and Zeileis, 2010) was used.

To fit either model function, we used the beta distribution parametrisation proposed in Ferrari and Cribari-Neto (2004); Cribari-Neto and Zeileis (2010). The 95% prediction interval in either case was defined with 2.5% and 97.5% quantiles of the beta distribution. To the best of our knowledge, beta regression — linear or nonlinear — has not yet been applied to model correlates of vaccine protection.

The data and R code used for this paper are freely available in a GitHub repository.

## B An approximation to the effectiveness model

### Complexities

That the population protection function (2) is defined over (distinct) distributions can complicate assessment of effectiveness predictions. Although predictions can be plotted against geometric mean titres (GMT), as in this paper, interpretation of the plots is not straightforward, since the underlying titre distributions are hidden. Technically, the model function is not a function of GMT: for instance, it could map the same GMT to multiple effectiveness values. Since we use empirical distributions, rather than, say, Normal distributions with a constant variance across variants in Khoury et al. (2021), our model function inputs do not reduce to two or three dimensions. Furthermore, a distribution’s shape could vary substantially between populations, which poses concerns about robustness of inferences. A distribution’s (geometric) mean, on the other hand, is usually more stable.

### A simplified model for effectiveness

To overcome these difficulties we define a simplified model as follows. All the titres per dose/variant combination for a vaccine are equal to the GMT of the corresponding sample from the Crick study. Then, the population protection function (2) coincides with the individual protection function (1). The simplified model assumes the latter function for vaccine effectiveness. Now the model function can be interpreted as a function of GMT only; see Figure 8.

**Figure 8:**
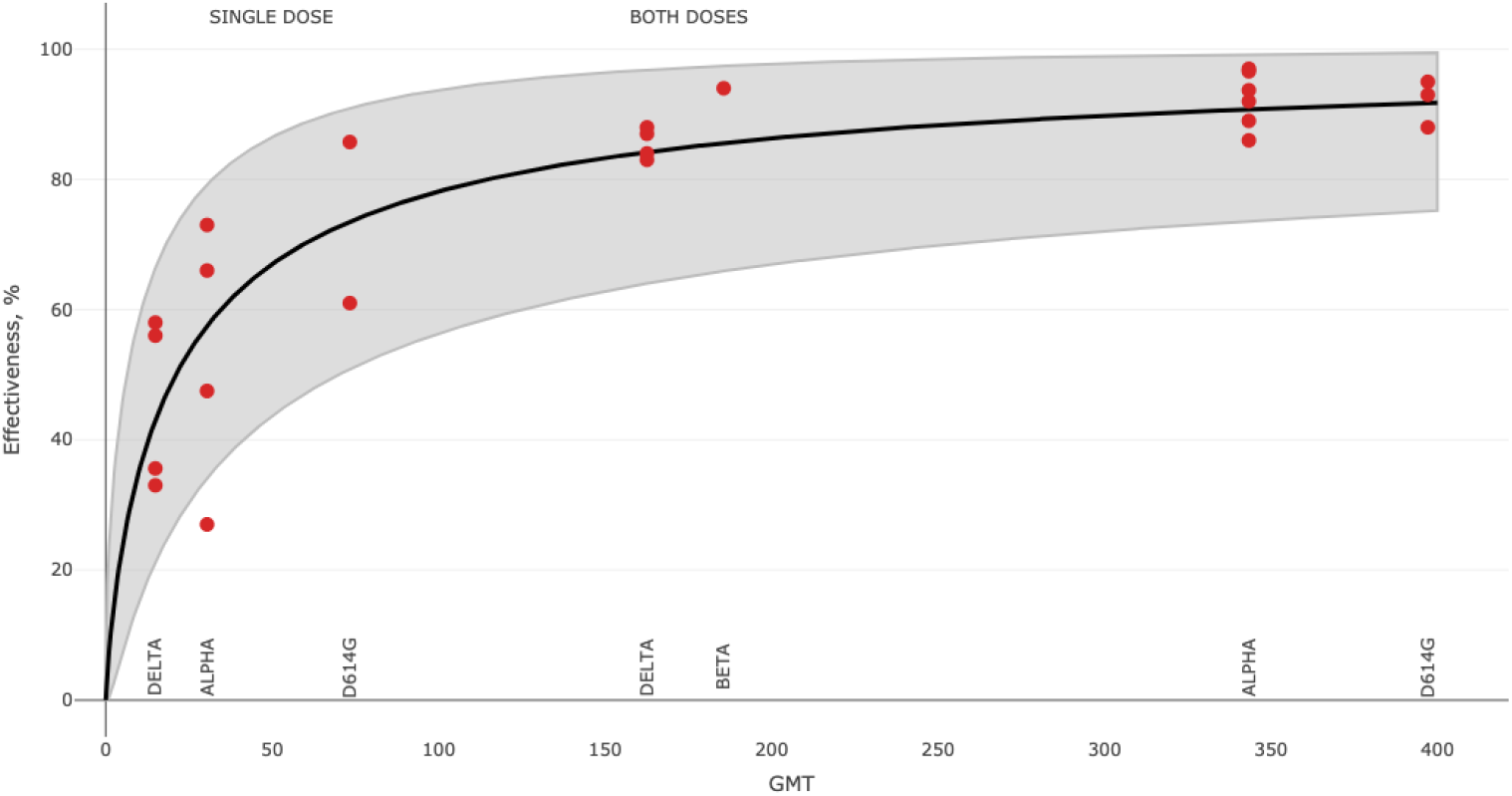
Simplified model fit to Comirnaty effectiveness observations, with 95% prediction bands shown.

Figure 9 shows that the two models and their prediction bands are close. This is particularly true after two doses for all the variants and after one dose for Alpha and D614G. We could therefore interpret the population protection function in Figure 1 as approximately equal to the simplified model function of GMT. Furthermore, predictions could be robust to the shape of log-titre distributions per variant, provided their GMTs are stable.

**Figure 9:**
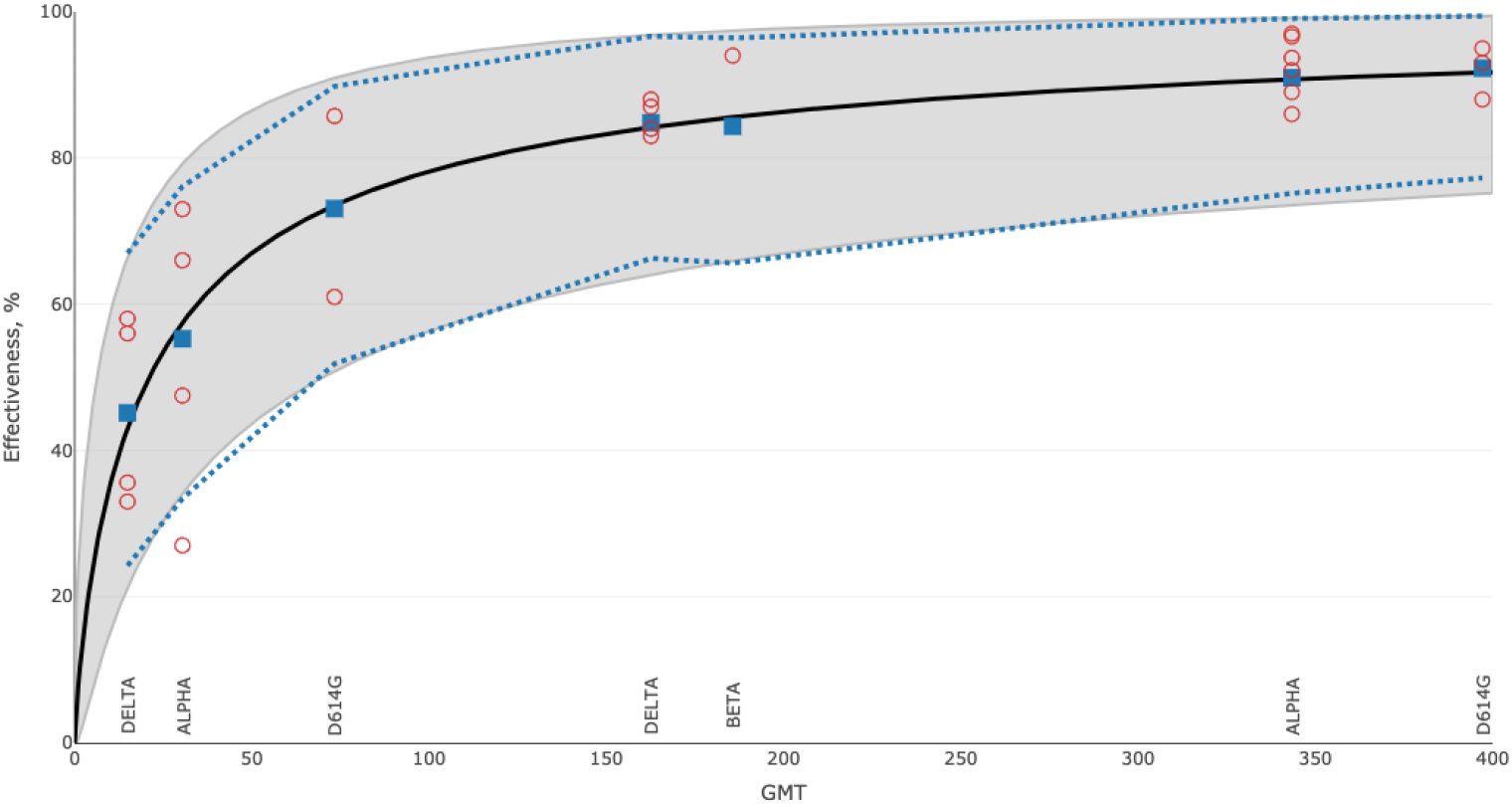
Simplified versus standard model fits for Comirnaty. Simplified fit and prediction bands are shown in a black line and grey shading. The standard model fit and prediction bands are shown in blue squares and dashed lines.

## C Neutralising titres versus age and other covariates

Some covariates can be strongly associated with immune response and thus matter for vaccination policies. The section considers dependence of neutralising titres on covariates for the Comirnaty vaccine.

The levels of Age covariate were set to the midpoints of five-year age bands in the Crick dataset. The Dosing Interval covariate for the two-dose cohort was dichotomised as “less than 40” or “more than 40” days between vaccine doses. The 40 days cut-off separated two distinct clusters in the data: at or near 21 versus at or near 63 days after the first dose.

### Two-dose cohort

A linear regression was fitted to log10 titres against the Delta variant (results for the other variants were similar). The *crude* fit used only Age, whereas the *multivariate* fit also used other covariates from the Crick data; see Table 1.

#### Categorical covariates

Dosing Interval and Prior Covid were the most significant categorical covariates in Table 1. Neutralising log-titres grouped according to these two covariates are shown in Figure 10. As mean ages per group were close — all within [42.7, 43.7] — we assumed that Age was not a confounder here. The figure confirms a well-known phenomenon that prior Covid-19 boosts antibody titres in vaccinees, compared to Covid-naive vaccinees. Our findings are in line with those in Payne et al. (2021) that larger dosing intervals for the Comirnaty vaccine increase antibody levels. In fact, our results suggest that Covid-naive vaccinees with a long interval could have higher titres than previously symptomatic vaccinees with a short interval. (Although note the small size, *N* = 17, of the latter subgroup.)

**Figure 10:**
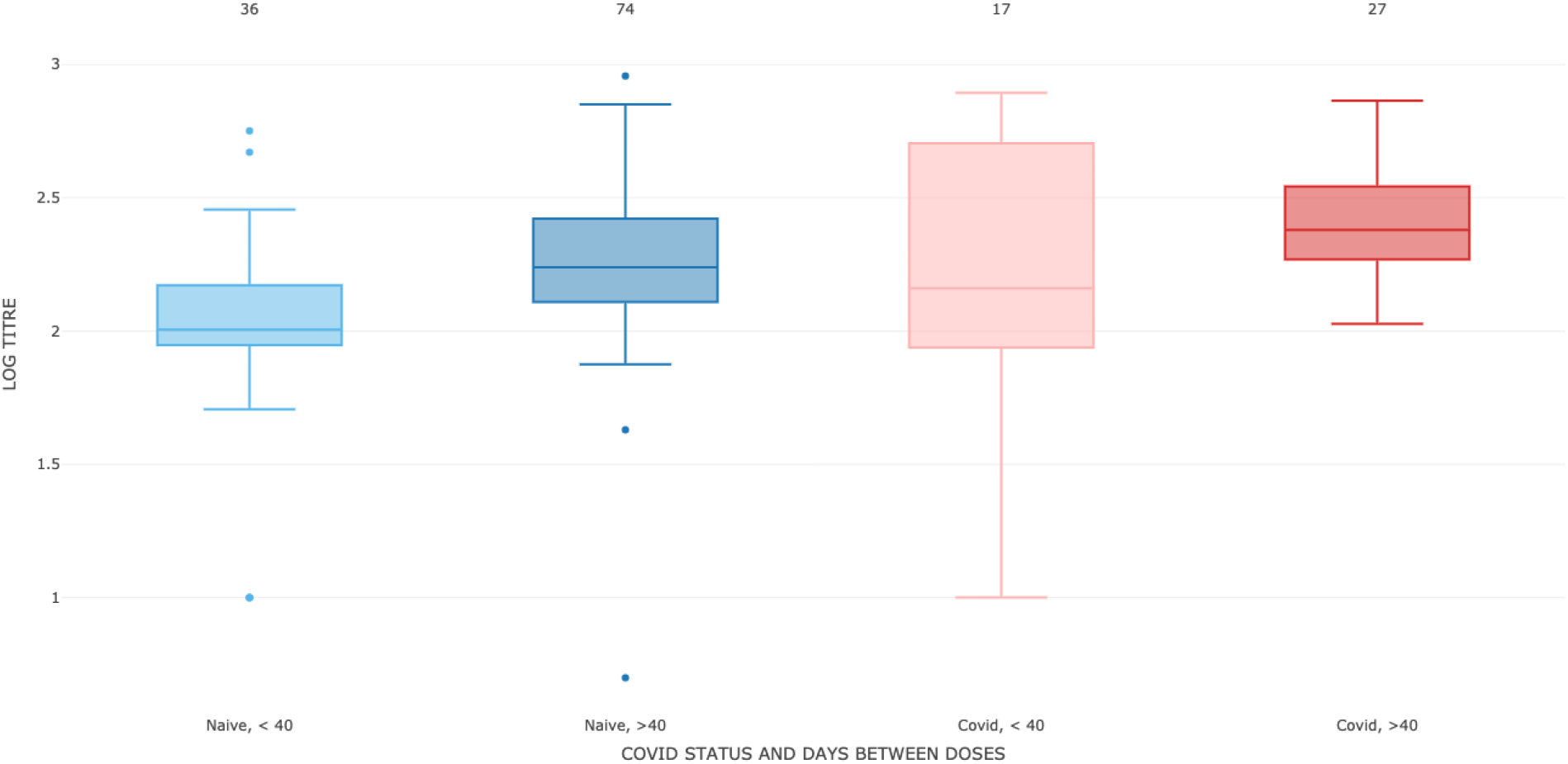
Two-dose Comirnaty cohort participants were grouped by Prior Covid-19 symptoms and Shorter versus Longer dosing interval. The log10 titres are shown for the Delta variant. The group sizes are indicated. The mean ages per group, from left to right, were 43.4, 42.7, 43.2 and 43.7.

#### Age covariate

We now associate log-titres with Age only, while treating other covariates as potential confounders. (This association, as modelled by the crude linear regression, is illustrated in Figure 11.) The Age coefficients in the crude and multivariate regressions differ by about 10%; see Table 1. As discussed in Chapter 11 of Kleinbaum et al. (1988), this difference may be inconsequential in some situations, in which case confounding could be ignored.

**Figure 11:**
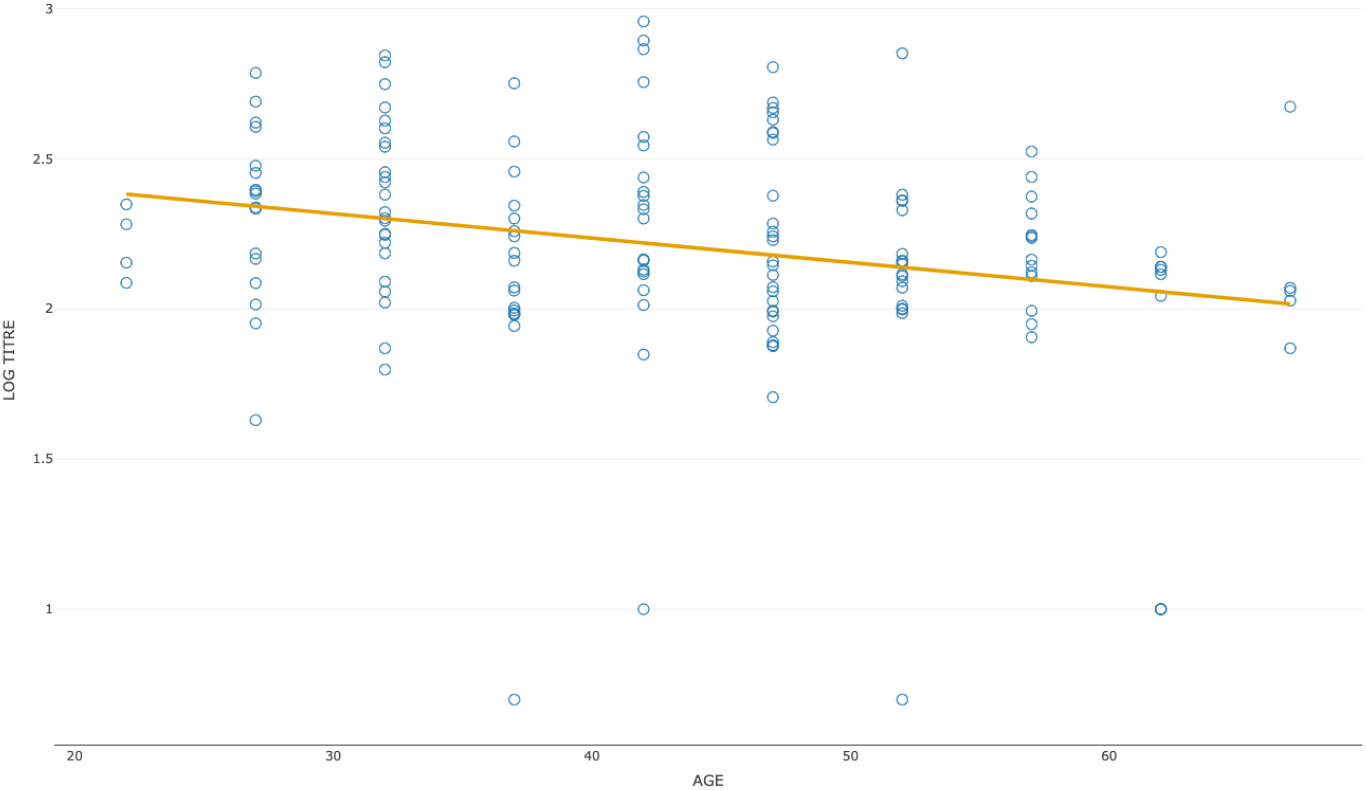
Log10 titres for Delta versus Age for the two-dose Comirnaty cohort. The fitted line corresponds to the crude linear regression in Table 1.

### Single-dose cohort

Linear regression results for the single-dose Comirnaty cohort are summarised in Table 2. Besides Age, only Prior Covid has a significant coefficient. As with two doses, the Age coefficients from the crude and multivariate regressions differed by about 10%. Note, however, that due to many titres below the limit of detection, the crude regression line is rather distant from the data; see Figure 12.

**Figure 12:**
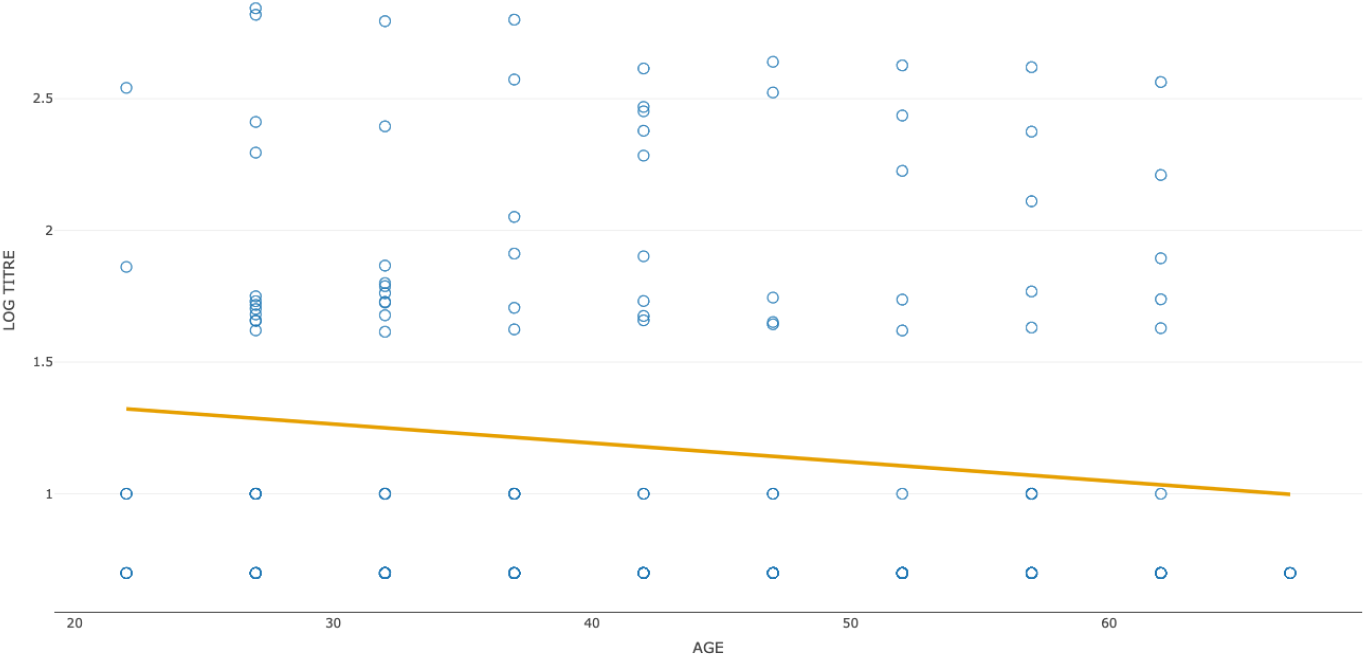
Log10 titres for Delta versus Age for the single-dose Comirnaty cohort. The fitted line corresponds to the crude linear regression in Table 2.

### Age groups

This paper mainly focuses on *below 35* and *50 and over* age groups; see, for example, Figure 2. Table 3 implies that these groups from the single-dose cohort are reasonably balanced with respect to Prior Covid, the significant confounder in Table 2. Likewise, Table 4 implies that these groups from the two-dose cohort are reasonably balanced with respect to Dosing Interval and Prior Covid, the significant confounders in Table 1. This balance and the close coefficients for Age in Tables 2 and 1 imply that the comparisons of these age groups per cohort are probably not strongly influenced by confounding.

**Table 3:** Relative subgroup sizes, in %, for the single-dose cohort

| Prior Covid | Age <35 | Age ≥ 50 | All Ages |
| --- | --- | --- | --- |
| N | 70.2 | 69.5 | 72.6 |
| Y | 29.8 | 30.5 | 27.4 |

**Table 4:** Relative subgroup sizes, in %, for the both-doses cohort

| Prior Covid | Dose Interval, days | Age <35 | Age $\geq 50$ | All Ages |
| --- | --- | --- | --- | --- |
| N | <40 | 22.7 | 22.2 | 23.4 |
| N | >40 | 47.7 | 48.9 | 48.1 |
| Y | <40 | 6.8 | 8.9 | 11.0 |
| Y | >40 | 22.7 | 20.0 | 17.5 |

Neutralising titres against Delta were compared per dose in younger (below 35), older (50 and over), and all-age participants in the Comirnaty stream. Non-parametric bootstrap was used to estimate the GMT per group and the GMT fold ratio for the younger versus older groups; see Figure 13. In almost all cases, the younger group had 1.6-2.3 times higher GMT than the older group did. The exception was the Beta variant after a single dose, possibly due to most individual titres being near or below the detection threshold; see Figure 7.

**Figure 13:**
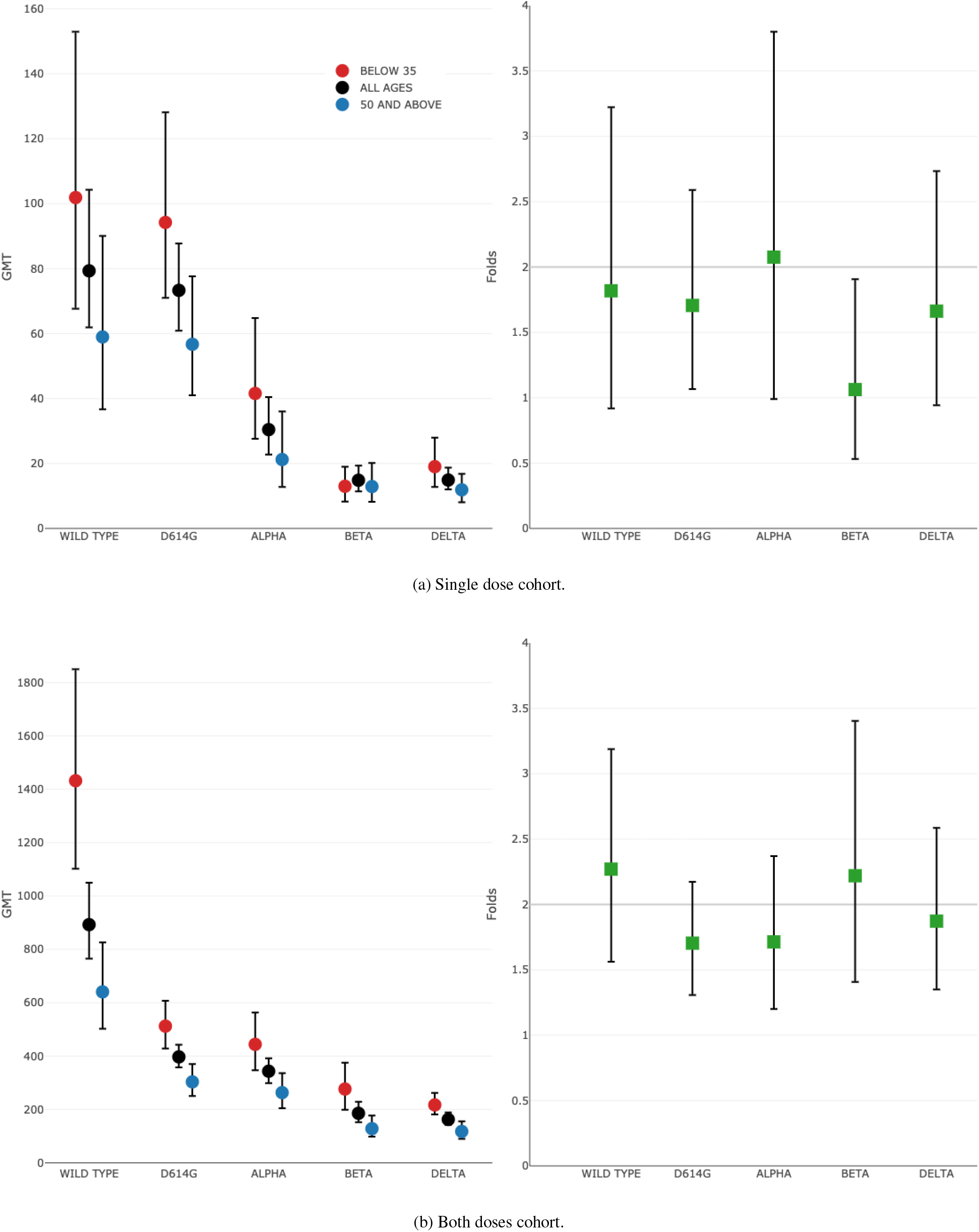
Geometric mean titres and their folds in age groups from the Comirnaty stream. The folds are for those below 35 versus those 50 and above. Also shown are 95% bootstrap confidence intervals. Data source: Wall et al. (2021b)

## D Population differences

### D.1 An example

The modelling in the main paper assumed that the titres distributions from the Crick study were representative of corresponding distributions in the effectiveness studies. To explore the impact of assumption violations due to population differences, we consider a simplified example for the Comirnaty vaccine.

Suppose that the “true population” in the effectiveness studies was the Comirnaty-vaccinated population in the Scotland study (Sheikh et al., 2021). We assumed Age as the only covariate for neutralising titres. (Neutralisation data from the actual Scotland population were not available.) The true “Scotland population” of titres was defined by using the age distribution, in Figure S2 of Sheikh et al. (2021), of Comirnaty vaccinees in Scotland to reweight the titres in the Comirnaty cohorts from the Crick study. As the Scotland study population was noticeably older than the Crick population, the constructed “true population” is noticeably different, according to Figure 14.

**Figure 14:**
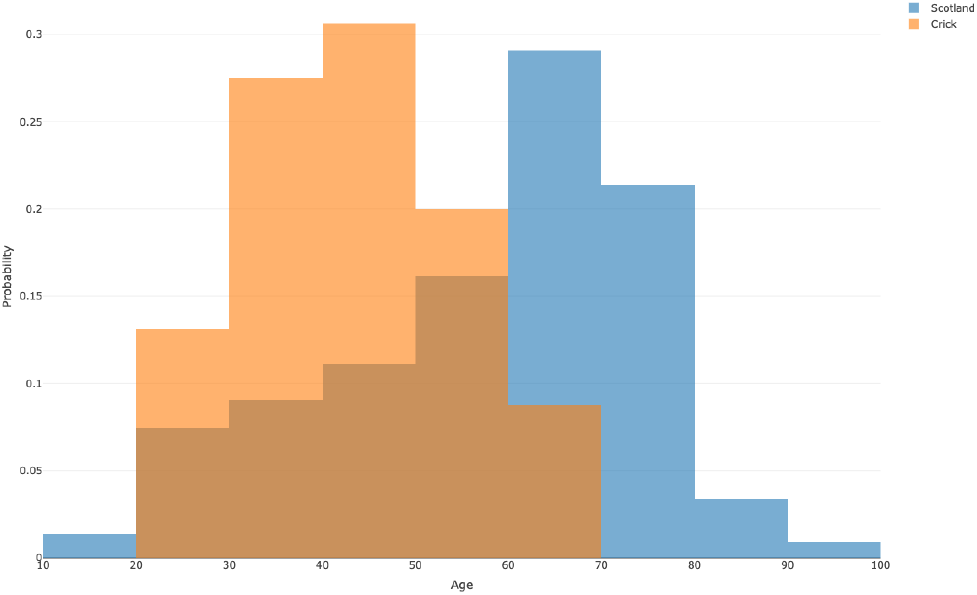
Age distribution for Comirnaty-vaccinated subjects in the Scotland study (Sheikh et al., 2021) versus that in the two-dose cohort in the Crick study (Wall et al., 2021b)

The effectiveness model was fitted using the “true” (weighted) and “original” (unweighted) titres; Figure 15. The weighted fit is shifted to the left, since lower titres, associated with older participants, had higher weights than in the original Crick data. Suppose that all the effectiveness observations indeed came from the Scotland study population, but the Crick study population were used in fitting. Then we would infer that higher than actual titres were needed for a given level of vaccine effectiveness and potentially underpredict it.

**Figure 15:**
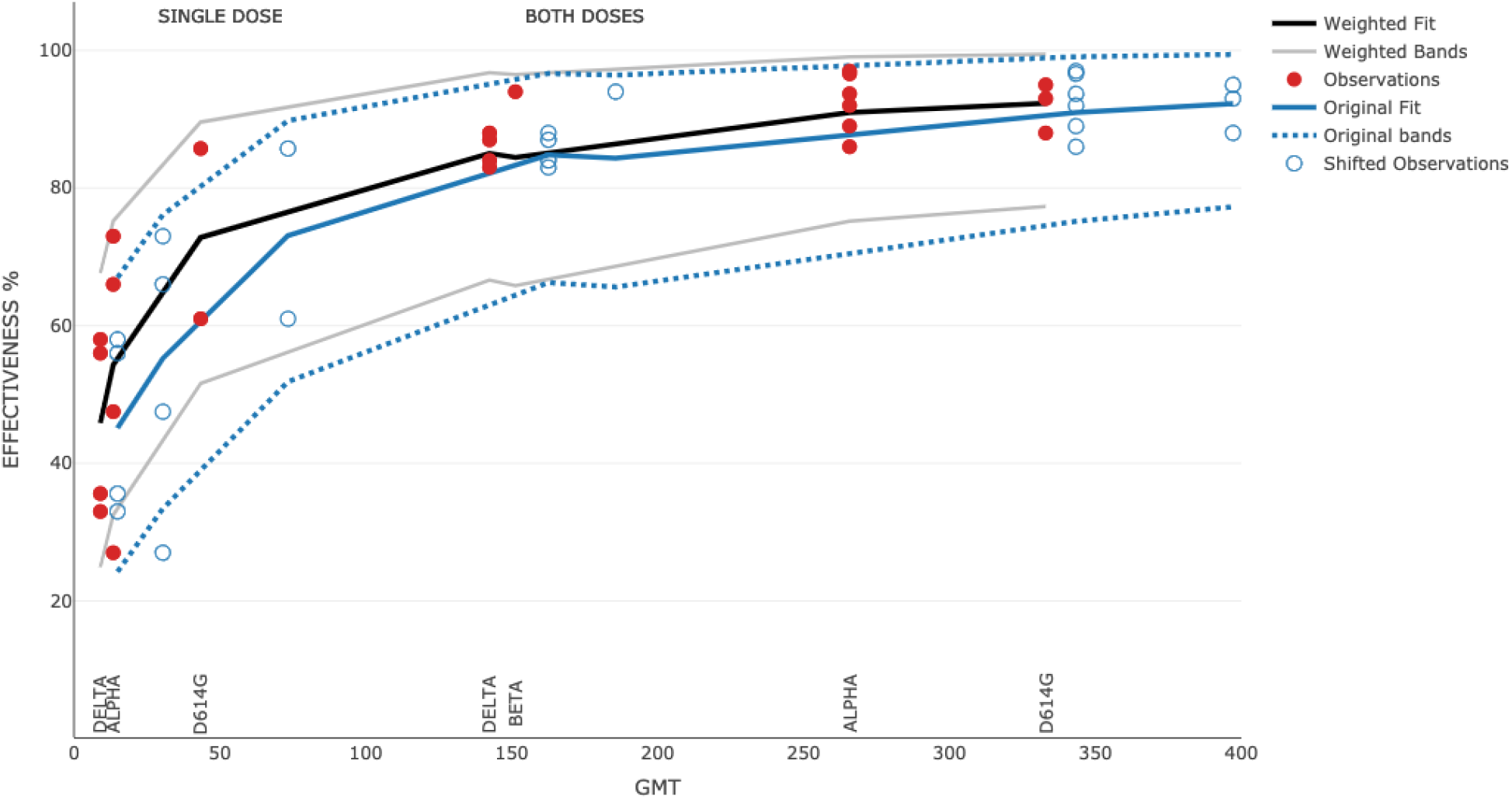
Model fits using age-weighted versus unweighted (original) neutralising titres from the Crick study with Comirnaty. The observations are plotted versus weighted and unweighted GMT per dose/variant. The variant labels are for weighted distributions.

While both weighted and unweighted model fits in Figure 15 appear reasonable, their predictions could differ substantially. Unless potential biases are heeded, a “good” model fit could be misleading.

### D.2 Statistical adjustments

The following adjustments could reduce biases due to population differences:

#### Iterative proportional fitting (aka data raking)

Neutralisation titres are iteratively weighted according to covariates so as to reflect participants in effectiveness studies more closely. The age-based reweighting in the previous section is in the spirit of this approach.

#### Propensity scores weighting

Weights can be defined to account for selection biases in relevant populations.

#### Subjects matching

Neutralisation study participants can be matched by covariates to their closest vaccinated subjects in an effectiveness study. Each effectiveness study would then provide a matched subset of participants. Effectiveness values would be re-estimated for these subsets and fit to the neutralisation titres per variant.

This matching could also proceed in the opposite direction. Suppose that enough vaccinated subjects are selected for a neutralisation study to cover relevant covariates in wider vaccinated populations. Each effectiveness study population could then be matched to a subset of the neutralisation study population. The effectiveness values would be fit to neutralisation data from these matched subsets.

#### Other approaches

Various other modelling approaches, such as sensitivity analyses (as illustrated below) and multimodel inference, could be also beneficial.

These adjustments rely on detailed information about both neutralisation and effectiveness studies populations. Such information is publicly available on (GitHub) for the Crick study but unfortunately not for effectiveness studies. Furthermore, some of the adjustments may require a specially designed trial. Therefore they were not investigated for this paper, but could be investigated in future.

### D.3 Equal multiples of titre distributions

Predictions from a neutralisation study would equal predictions per dose/variant from the true population under the following “proportionality” condition. Each true titre distribution per dose/variant can be obtained by multiplying the corresponding neutralisation study distribution by some constant *κ >* 0. This *κ* must be the same for all dose/variant combinations but need not be known.

The condition can be seen if we consider log *x*_*i*_ and log *κx*_*i*_, denoting the log-titre for subject *i* in the neutralisation study and person *i* in the true population, respectively. (To simplify the indexing, suppose that both populations have equal size *N*.) The corresponding terms in model (2) are *α*+*β* log *x*_*i*_ and *α*+log *κ*+*β* log *x*_*i*_, which differ by a constant independent from the titre. Consequently, a model fitting neutralisation study titres to effectiveness observations would produce exactly the same predictions as a model fitting true titres to the same observations; see Figure 16.

**Figure 16:**
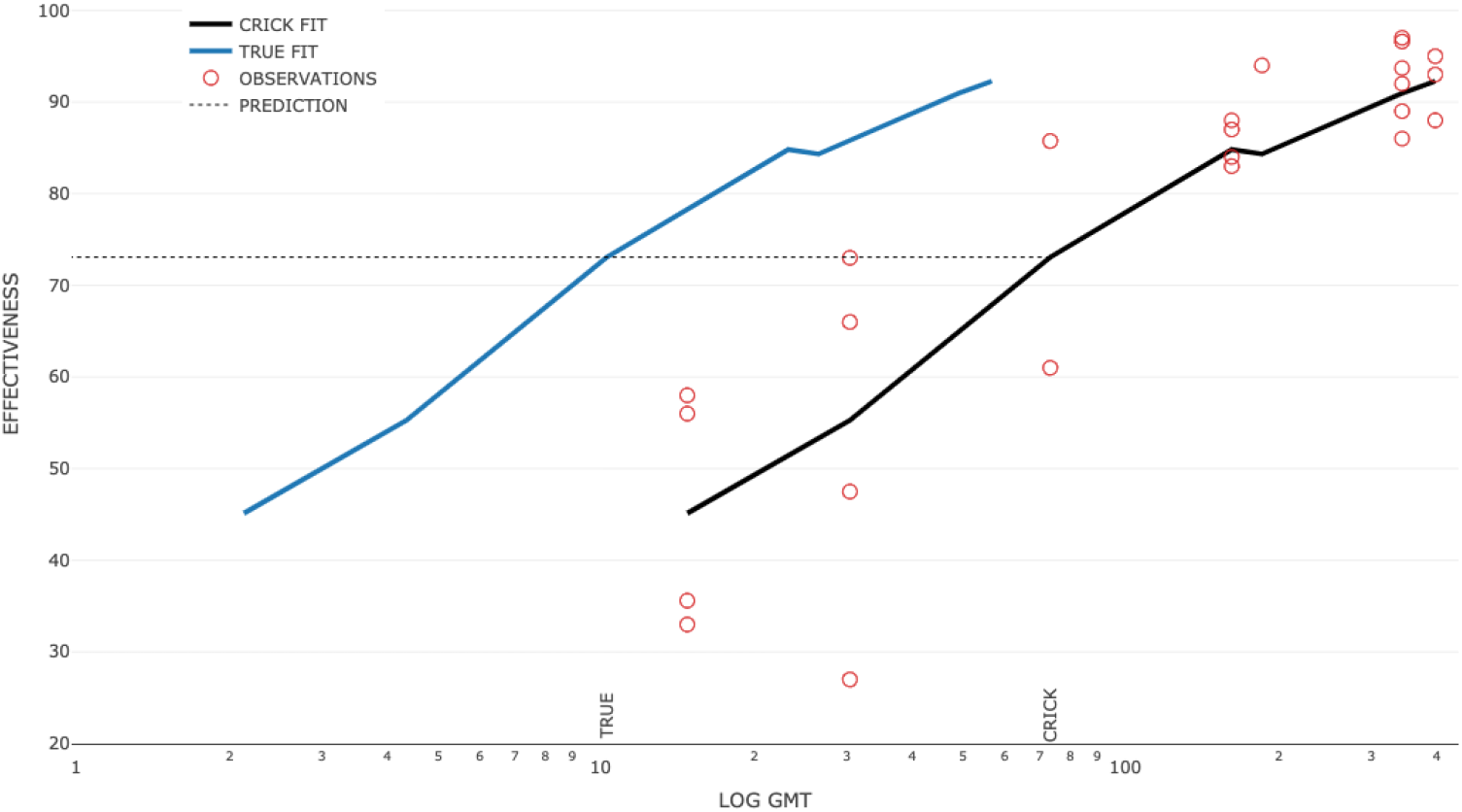
True distributions of titres are a constant multiple of Crick distributions

Although this condition is unlikely to hold at the outset, it could hold approximately after population adjustments. It also implies that titre distributions need not be matched perfectly, but only to make them approximately proportional. Conveniently, the actual value of *κ >* 0 need not be known.

### D.4 A sensitivity investigation

We consider an example where “true” and “neutralisation study” titre distributions are mismatched. Suppose effectiveness predictions against the Delta variant are required for the Comirnaty vaccine, as in Section 1.2.

To define a “true” titre distribution per dose/variant combination *j* we start with the corresponding distribution from the Crick study. We next apply the distribution according to the age distribution from the Scotland study in Sheikh et al. (2021), as done in Section D.1. Each distribution *j* is multiplied by *κ*_*j*_ *>* 0, with these constants possibly different between dose/variant combinations. The resulting “true” distributions have a different shape from corresponding “Crick” distributions and violate the proportionality condition.

The constant *κ*_*j*_ was simulated per dose/variant combination *j* under the following scenarios:

**DOWN** All titre distributions decrease (i.e., shift to the left), with *κ*_*j*_ drawn from [0.25, 1]

**Down/up** Distributions are more likely to decrease than to increase (i.e., shift to the right). The *κ*_*j*_ is drawn from [0.5, 1], with probability 0.67 and from [1, 2] with probability 0.33.

**Down/Up** Distributions are equally likely to decrease or increase. The value *κ*_*j*_ is drawn from [0.5, 1] and from [1, 2] with equal probabilities

**Up/down** Distributions are more likely to increase, with *κ*_*j*_ drawn from [0.5, 1] with probability 0.33 and from [1, 2] with probability 0.67.

**UP** All distributions increase, with *κ*_*j*_ drawn from [1, 4].

The algorithm randomly chose an interval, when required by the scenario, and then drew *κ*_*j*_ from a uniform distribution over the interval. The simulation ensured that the GMT after a first dose for a particular variant was less than that after two doses. For each scenario, 1000 vectors of *κ*_*j*_ were simulated, with 5000 vectors in total. Hence, each vector of *κ*_*j*_ defines a set of true titre distributions. Each of these sets was used to fit the model to the Comirnaty effectiveness observations, *including* those for the Delta variant.

The predicted effectiveness values against Delta, after one and two doses were set as the true effectiveness values. We note their relatively small variability despite the wider ranges used for *κ*_*j*_ values. (In every scenario, the *κ*_*j*_ values could differ by up to four folds.) According to Figure 17, the smallest lower quartile of predictions per scenario is 82.1%, and the largest, 87.1%, for two doses, and 44.2% and 50.3% for one dose.

**Figure 17:**
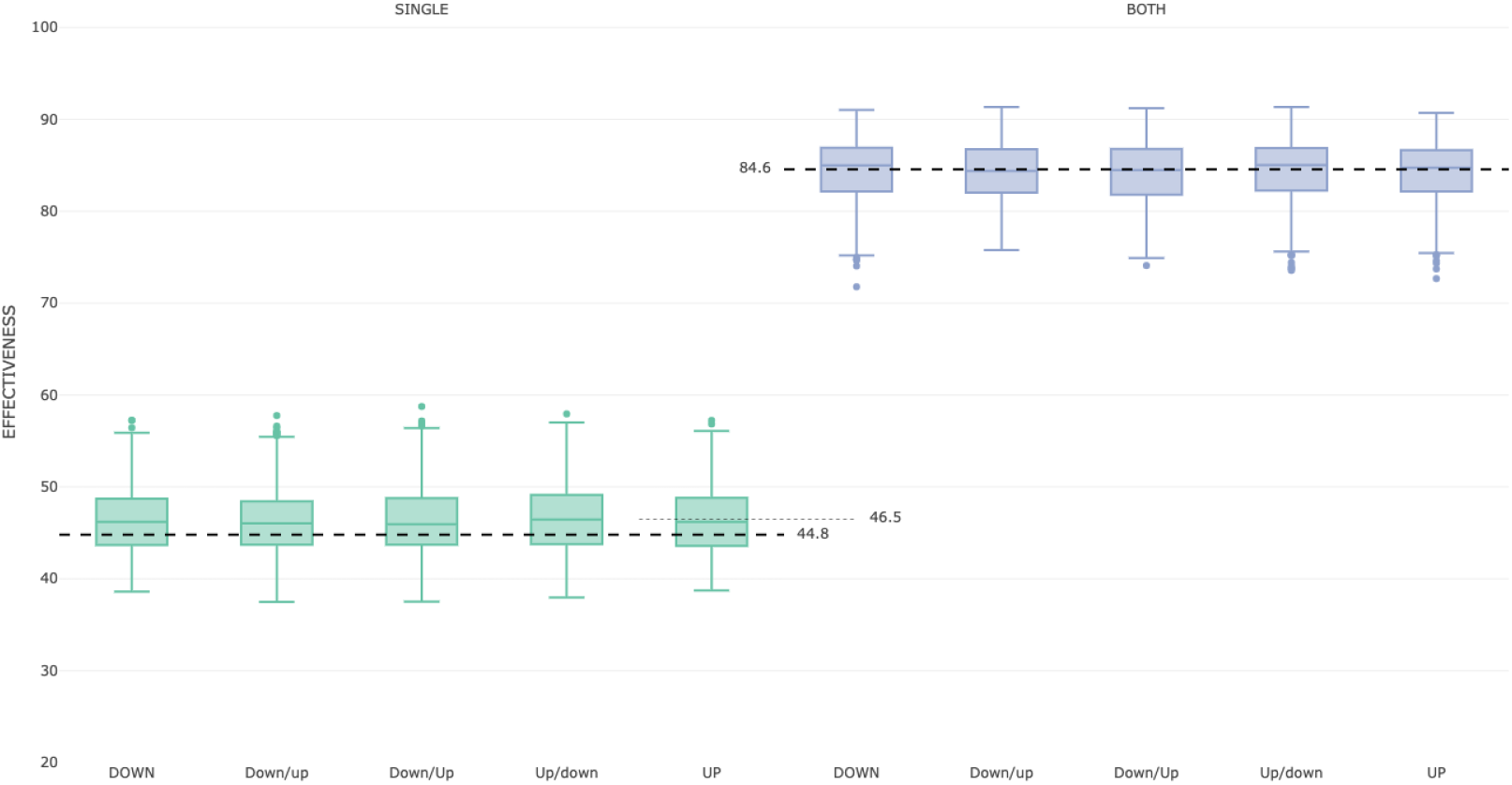
Robustness of effectiveness predictions against Delta for the Comirnaty vaccine. Crick study population versus Shifted Age-Weighted “true” distributions per dose. Box plots show effectiveness values evaluated for the Delta variant at a model fitting all effectiveness observations to the simulated true titres. The thick dashed lines show predictions from the unweighted Crick neutralisation data fitted to the effectiveness observations without Delta. The thin dashed line shows prediction using age-weighted Crick data. (After both doses, weighted and unweighted predictions were almost equal.)

Predictions from the model fit to original Crick data were used as a baseline. The same model fit was used in Section 1.2 to predict effectiveness against Delta; see Figures 3 and 4. This fit did not use data on the Delta variant.

The baseline predictions were compared with simulated “true” predictions per scenario; see Figure 17. The medians of true predictions (per scenario) after one dose were in [46.6, 47.2], compared to 44.8 for the baseline prediction. After two doses, they were in [84.6, 85.3] compared to 84.6. The baseline predictions had a small bias after two doses and a slightly larger bias after one dose.

To explore the effect of population matching we compared the simulated “true” predictions with predictions from age-weighted Crick distributions. The age-weighted predictions, 46.5 and 84.9, for one and two doses, respectively, were both closer to the corresponding medians of simulated predictions, which illustrates the usefulness of population adjustments.

### D.5 Cautious prediction

Suppose that the Crick study population — possibly after an adjustment — has similar or slightly higher neutralisation titres than a true population does. There could still remain uncertainty about how closely the populations match and how well the true titres are approximated.

One way to mitigate this uncertainty is to include a safety margin for effectiveness predictions, as follows. First we select a sub-population of the Crick participants with higher, on average, neutralisation titres. Here we simply choose all the participants aged under 35. We next fit their neutralisation titres to the effectiveness observations; see Figure 18. The resulting fit — based on higher titres — is to the right of the all-age fit. To predict effectiveness against a new variant, for example, we would collect neutralisation titres against it in the all-age population. To make a cautious prediction we input these titres into the under-35 model.

**Figure 18:**
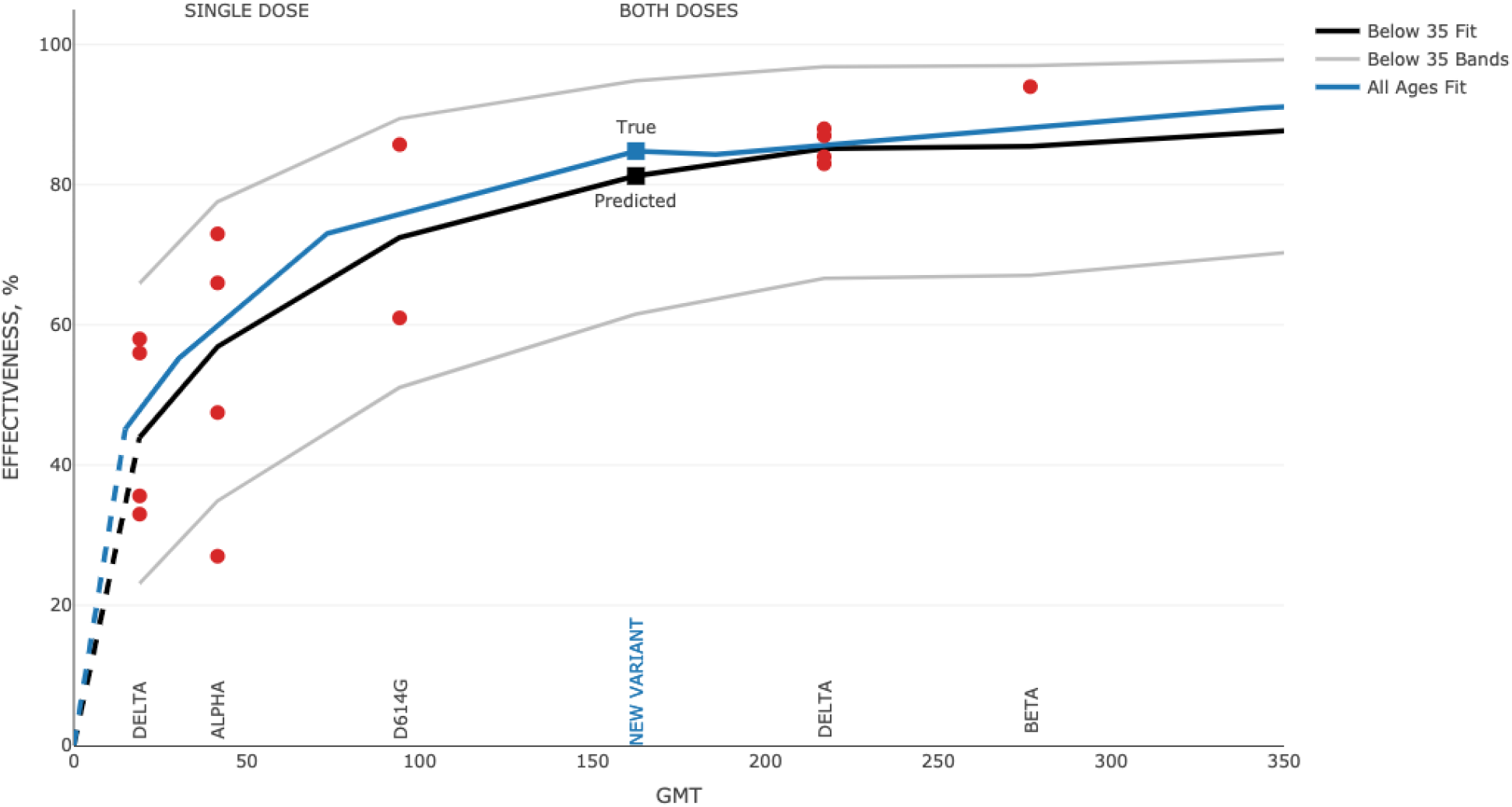
Cautious prediction for Comirnaty. Observations were fitted to the below 35 age group; the all-ages fit is shown for comparison. Effectiveness for a new variant was predicted for all-age titres but assuming the below-35 fit.

In a numerical example, illustrated in Figure 18, the cautious prediction, 81.3%, is 3.5% less than 84.8%, the all-age fit. By using the cautious prediction we can infer that the all-age effectiveness would be at least 81.3% and likely higher. By extension, we infer that the effectiveness in the true population would be approximately equal to and possibly higher than 81.3%.

This cautious approach could be practically beneficial. It requires just enough information to assume that a true population has similar or lower titres than a neutralisation study population does. The true distribution is not required, and the impact of population mismatch can be reduced. If a cautious prediction, say, of 85% effectiveness after a half-booster, is still high enough in practice, then a sound decision about the booster could be made. The cautious approach could be also useful when combining effectiveness data from disparate populations in different effectiveness studies. Excessive caution, however, is undesirable, as it may, for instance, lead to boosting too early.

We note that the unweighted fit in Figure 15 could enable cautious predictions for the assumed true population. The same conclusion can be made from Figure 17, especially for the first dose. We could therefore use the Crick study population as-is or apply the approach in this section for an extra margin of safety.

## E Extended Data

### Comirnaty vaccine

We performed a literature search for studies of symptomatic effectiveness (and efficacy) with the Comirnaty vaccine. We sought studies either specifying the assumed SARS-CoV-2 variant or for which the predominant variant could be inferred from the study dates and location. We identified 24 effectiveness (and efficacy) observations in all-age groups from eight studies; see Table 5.

**Table 5:** Studies reporting symptomatic effectiveness estimates for the Comirnaty vaccine in all-age populations. Eff. stands for efficacy for Study 1, and effectiveness for the other studies.

| Study | Dose | Days | Variant | Country | Eff. | Confidence | Reference |
| --- | --- | --- | --- | --- | --- | --- | --- |
| 1 | 1 | 14 | D614G | US | 85.7 | (59.3, 97.5) | FDA (2020) |
|  | 2 | 7 | D614G | US | 95.0 | (90.0, 97.9) | Polack et al. (2020) |
| 2 | 1 | 14 | D614G | Canada | 61.0 | (54.0, 68.0) | Nasreen et al. (2021) |
|  | 1 | 14 | Alpha | Canada | 66.0 | (64.0, 68.0) |  |
|  | 1 | 14 | Delta | Canada | 56.0 | (45.0, 64.0) |  |
|  | 2 | 7 | D614G | Canada | 93.0 | (88.0, 96.0) |  |
|  | 2 | 7 | Alpha | Canada | 89.0 | (86.0, 91.0) |  |
|  | 2 | 7 | Delta | Canada | 87.0 | (64.0, 95.0) |  |
| 3 | 1 | 28 | Alpha | Scotland | 27.0 | (13.0, 39.0) | Sheikh et al. (2021) |
|  | 1 | 28 | Delta | Scotland | 33.0 | (15.0, 47.0) |  |
|  | 2 | 14 | Alpha | Scotland | 92.0 | (88.0, 94.0) |  |
|  | 2 | 14 | Delta | Scotland | 83.0 | (78.0, 87.0) |  |
| 4 | 1 | 21 | Alpha | England A | 47.5 | (41.6, 52.8) | Bernal et al. (2021) |
|  | 1 | 21 | Delta | England A | 35.6 | (22.7, 46.4) |  |
|  | 2 | 14 | Alpha | England A | 93.7 | (91.6, 95.3) |  |
|  | 2 | 14 | Delta | England A | 88.0 | (85.3, 90.1) |  |
| 5 | 2 | 7 | D614G | France | 88.0 | (81.0, 92.0) | Charmet et al. (2021) |
|  | 2 | 7 | Alpha | France | 86.0 | (81.0, 90.0) |  |
| 6 | 2 | 7 | Alpha | Israel A | 96.6 | (95.8, 97.2) | Haas et al. (2021) |
| 7 | 2 | 14 | Beta | Israel B | 94.0 | (90.0, 98.0) | Mor et al. (2021) |
| 8 | 1 | 21 | Alpha | England B | 73.0 | (68.0, 76.0) | Pouwels et al. (2021) |
|  | 2 | 14 | Alpha | England B | 97.0 | (96.0, 98.0) |  |
|  | 1 | 21 | Delta | England B | 58.0 | (51.0, 64.0) |  |
|  | 2 | 14 | Delta | England B | 84.0 | (82.0, 86.0) |  |

The first study is the Pfizer-BioNTech Phase 2/3 vaccine trial mostly conducted in the US (130 out of 152 study sites). The estimate of symptomatic efficacy after two vaccine doses was given in Polack et al. (2020) for the study period between May and November 2020. During that time, more than 90% of the SARS-CoV-2 infections in the US, and similarly world-wide, had the D614G mutation Korber et al. (2020). Hence, we assigned D614G to the efficacy estimates from this study.

Following Skowronski and Serres (2021), we also calculated the vaccine efficacy 14 days after the first dose, by using the revised efficacy data in the FDA submission (FDA, 2020). The estimate was made using the Bayesian approach used by Pfizer investigators to estimate efficacy after two doses (FDA, 2020).

We included seven observational studies as follows: one study in Canada (Nasreen et al., 2021), two in England (Bernal et al., 2021; Pouwels et al., 2021); one in France (Charmet et al., 2021), two in Israel (Mor et al., 2021; Haas et al., 2021) and one in Scotland (Sheikh et al., 2021). Note that (Charmet et al., 2021) reported combined effectiveness estimates for Pfizer-BioNTech and Moderna mRNA vaccines. We included this study as it stated that 87% of the participants had the former vaccine.

### Vaxzevria vaccine

A literature search was performed for studies reporting symptomatic effectiveness (and efficacy) for the Vaxzevria vaccine. The studies were sought that either specified the assumed SARS-CoV-2 variant or for which the predominant variant could be inferred from the study dates and location. We included the estimates of symptomatic effectiveness from Emary et al. (2021) who re-analysed the vaccine clinical trial (Ramasamy et al., 2020) to compute effectiveness against the Alpha variant and against an ancestral type, which we inferred to be D614G. We also included estimates from four observational studies with the vaccine post-authorisation: one in Canada (Nasreen et al., 2021); two in England (Bernal et al., 2021; Pouwels et al., 2021); and one in Scotland (Sheikh et al., 2021). Altogether there were 17 observations for D614G, Alpha and Delta variants in all-age populations after one or two doses; see Table 6. The study in Madhi et al. (2021) estimated the effectiveness of two doses of Vaxzevria against the Beta (B.1.351) as 10.4% (−76.8, 54.8). This estimate was identified as an outlier and was not used in model fitting in this paper.

**Table 6:**
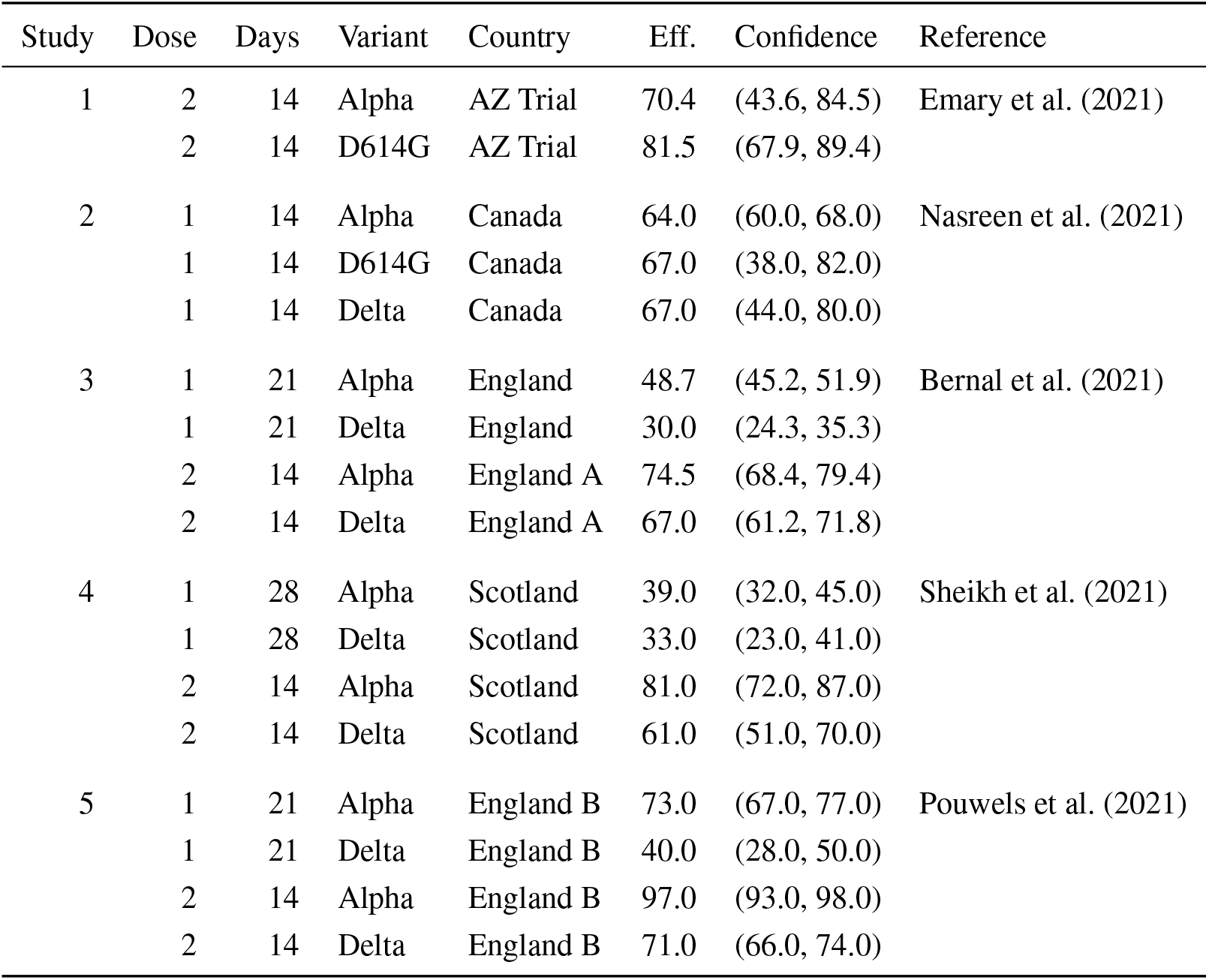
Studies reporting symptomatic effectiveness estimates for the Vaxzevria vaccine in all-age populations. Eff. stands for efficacy for Study 1, and effectiveness for the other studies.

